# Collective interactions, human mobility and viral evolution shaped the SARS-CoV-2 transmission in Mainland China

**DOI:** 10.64898/2025.12.17.25342513

**Authors:** Dong Wang, Yi Wang, Oswaldo Gressani, Jiaqi Chen, Yuxin Tao, Hao Wang, Shanshan Li, Dongxuan Chen, Eric H. Y. Lau, Yi Zhao, Peng Wu, Qingpeng Zhang, Benjamin J. Cowling, Sheikh Taslim Ali

## Abstract

Collective interaction of individuals in various settings is crucial for exposure to infections, encompassing complex viral interplay and amplifying infectious risk through phenomena such as social reinforcement, clustering and superspreading events, during the COVID-19 pandemic. However, standard epidemic models often inadequately capture such heterogeneity, overlooking the higher-order social structural. Spatiotemporal variation in transmission, an essential feature of the pandemic, remains poorly quantified at various scales, particularly in integrating high-resolution data streams and complex network approaches. We introduced a higher-order simplicial model that unifies human mobility data, genetic diversity and antigenic drift to systematically investigate the role of social reinforcement, spatiotemporal variation and genetic mutations in SARS-CoV-2 transmission. We found a median of 5.3% to 14.4% of infections across provinces were attributed to social reinforcement, while cluster heterogeneity contributed to a median of 17%–71% increase in susceptibility. Multiple viral interactions elevated transmissibility by 68%–70% across the periods of dominant variants. The reconstructed transmission networks underscored distinct spatiotemporal variation, with dynamic critical locations, varying mobility patterns, and evolving geographic cluster structures, by assessing complex networks. The influence of human mobility was found to be positive on transmission, effective distance was negatively associated with infection risks, while greater genetic diversity and antigenic drift were linked to higher susceptibility and transmissibility. Our proposed data-driven higher-order framework could help us to understand epidemics better by accounting the role of collective interactions, population mobility, and genetic mutation in transmission, which could inform the targeted interventions to mitigate SARS-CoV-2 and other respiratory pathogens.

---

The COVID-19 pandemic, caused by the SARS-CoV-2 virus ^1,2^, has posed unprecedented challenges on a global scale ^3^, underscoring the imperative for a comprehensive understanding of the dynamics and underlying transmission characteristics ^4,5^. Several studies have highlighted the pivotal role of superspreading and clustering events in shaping the transmission dynamics of SARS-CoV-2 ^6–8^. Social reinforcement ^9–12^, a phenomena that an individual behavior is influenced by collective interactions in groups such as households, workplaces, and restaurants, has emerged as a critical determinant of disease transmission, could occur not only after infection but also beforehand ^13–15^. This effect is particularly salient for SARS-CoV-2, a respiratory pathogen transmissible via aerosols^16^. Recent evidence further indicated that when multiple strains co-circulate in a community, they may involve in interaction, mutual competition, and undergo significant mutations, potentially resulting in novel strains with more infectiousness or immune evading properties ^17,18^. Incorporating the time-varying information on genetic mutation in epidemiological models is important to elucidating how viral evolution contributes to the transmission dynamics and outbreak trajectories ^19–22,23^. Standard epidemic models typically assume a homogeneous, linear, pairwise relationship between the number of infectious contacts and the risk of infection for susceptible individuals, but often fail to capture the complex heterogeneity introduced by social reinforcement within collective interactions ^9,10,13,24,25^. Accordingly, a higher-order modeling framework that integrates genetic information is warranted to elucidate the influence of social reinforcement and genetic dynamics embedded within collective interactions (human behaviors) on infectious disease transmission ^23,26–29^.

Moreover, pronounced spatiotemporal variation in transmission dynamics has been a defining features of the COVID-19 pandemic ^2,30^, particularly in large and diverse countries such as China, where social structures and mobility patterns differ substantially across provinces ^31–33^. Transmission patterns have demonstrated significant temporal variability, underscoring the time-varying shift in community transmission ^34^. Systematic analysis of spatiotemporal transmission variation facilitates the identification of high-risk regions at sub-national scale during specific periods and enables proactive implementation of targeted mitigation and control strategies ^32,34,35^. However, attempts to characterize long-term, nationwide, spatiotemporal transmission dynamics, leveraging high-resolution data and network-based epidemiological modeling analysis remain limited, underscoring potential scopes of further investigation ^32,34,36^.

Along with spatiotemporal variation in transmission risk, heterogeneity in human social reinforcement is fundamental for understanding disease dynamics. These key factors account for the intricate interplay of social interactions, viral co-circulation and population mobility were barely assessed for their impact in shaping transmission dynamics ^2,13,14,24,26,32,35^. To quantify the effects of social reinforcement on SARS-CoV-2 transmission and its spatiotemporal variation on outbreak patterns, we proposed a spatial higher-order mathematical modeling framework combined with complex network analysis, leveraging high-resolution surveillance data on epidemiology, human mobility and viral evolution across mainland China. Integrating social reinforcement within epidemiological models could provide critical insights into the challenges of controlling SARS-CoV-2 within collective settings. Furthermore, spatiotemporal analysis of transmission networks could strengthen our understanding of the mechanisms driving pandemic spread, which could inform for effective public health interventions plan. Our general higher-order modeling framework underscored the necessity of incorporating social reinforcement, genetic signals in the co-circulating dynamics, and human mobility into epidemiological models, offering a practical perspective for the prevention and mitigation of the burden of SARS-CoV-2 and other respiratory pathogens under heterogeneous population settings.

## Higher-order modelling with social reinforcement

We integrated the collective interaction with its impact as social reinforcement into our modeling framework to quantify its contribution to force of infection (FoI) for less or non-homogeneous population ^9,10^. To illustrate this, we constructed the higher-order model by incorporating social reinforcement against the standard pairwise model as illustrated in Fig. 1a-b. In the standard model, the FoI was expressed as *λ*(*t*)=*β*(*t*)*I*(*t*), where *β*(*t*) denoted the time-varying transmission rates and *I*(*t*) is the infection cases at time 𝑡. By incorporating nonlinear dynamics arising from collective interactions, the higher-order formulation extends the FoI to λ(*t*)=*β*(*t*)*I*(*t*)+*v*(*t*)*β*(*t*)*I*(*t*)^2^ , where *v(t)=*𝑣*_j_*_0_ (𝑡)𝑒^𝛼1\**div*(*t*)^ quantifies the transmissibility modulated by social reinforcement and genetic diversity. Here, 𝑣*_j_*_0_ (𝑡) represents the baseline impact of social reinforcement for dominant variant of concern 𝑗, 𝑑𝑖𝑣(𝑡) denotes genetic diversity at time *t* and 𝛼_+_ is coefficient reflecting its impact. This modeling framework transcends the limitations of traditional pairwise interaction paradigms by explicitly encoding group-level, collective transmission via social reinforcement ^9,10^. Within this higher-order modeling paradigm, social structures were assimilated as simplicial complexes, with environments such as households, workplaces, restaurants, and social gatherings characterized as higher-dimensional simplices ^24^. This formulation enabled an explicit characterization of collective interactions along with pairwise interactions, thereby overcoming the intrinsic limitations of traditional standard pairwise contact models.

**Figure 1.**
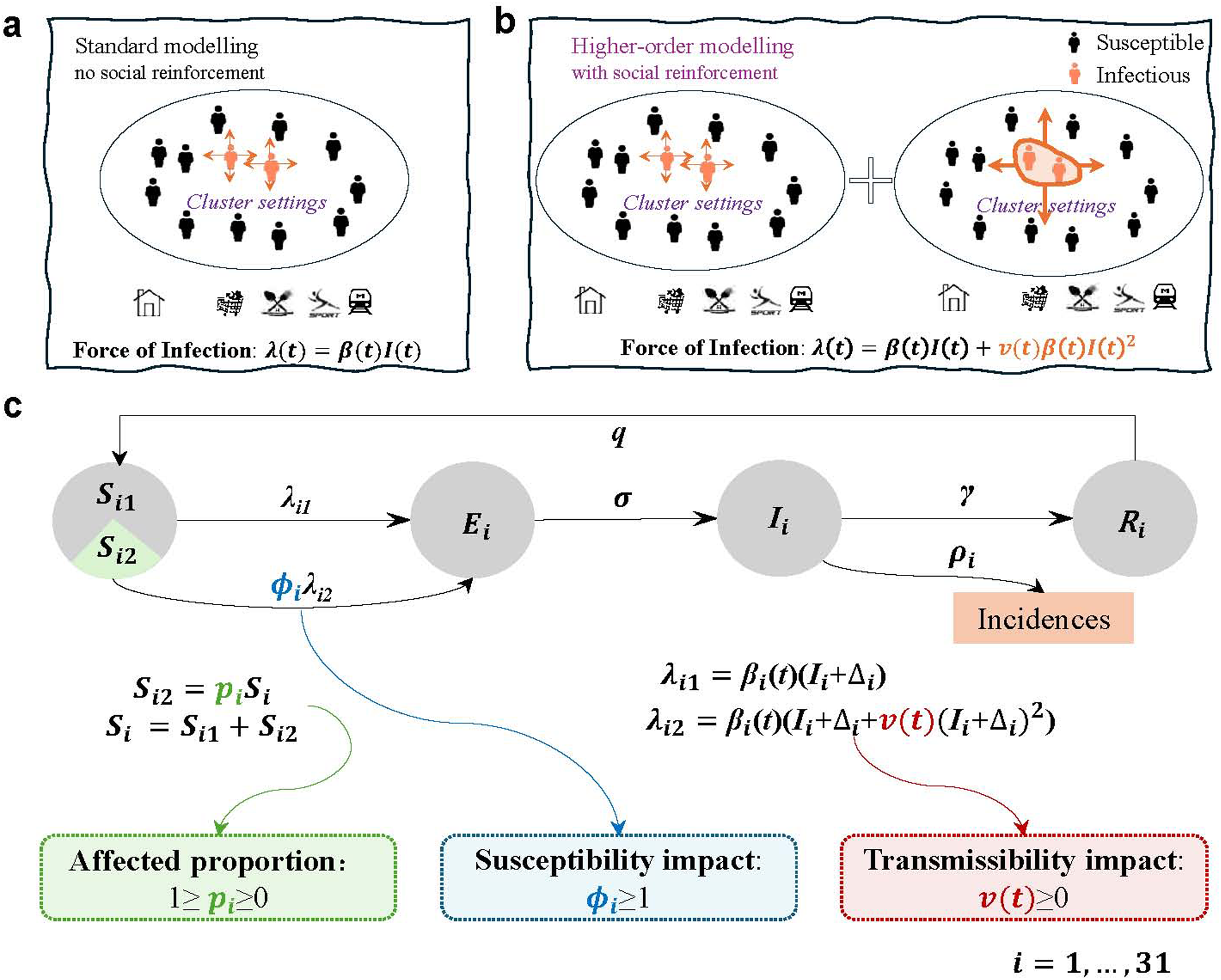
Epidemic modelling with and without social reinforcement, and higher-order modelling schematic representation under SEIR framework. (**a)** Schematic of the standard epidemiological model without social reinforcement, illustrating baseline infection risk. (**b)** Higher-order model incorporating social reinforcement, which modifies infection risk within cluster settings. The infection risk includes both the standard one *β*(*t*)*I*(*t*) and another one *β(t)I(t)^2^* from social reinforcements, where the formula *I*(*t*)^2^ in population, the level is equivalent to two infections in a cluster at the individual level. Susceptible and infected individuals are represented in black and red, respectively. **(c)** Schematic representation of SARS-CoV-2 transmission model under a simple SEIR framework, where individuals progress along *S_i_*→*E_i_*→*I_i_*→*R_i_*→*S_i_* at per capita rates *λ*, *σ*, and *γ* for province *i*, respectively. The progression of individuals infected not under social reinforcement is tracked via classes *S_i_*_1_ and *S_i_*_2_ , where *p*(*t*) is the proportion of infected individuals under social reinforcements. *ϕ_i_*>1 captures the enhanced susceptibility arising from collective interactions, as determined by cluster heterogeneity. *v*(*t*)>0 represents the increased transmissibility conferred by the concurrent presence of two infectors within a cluster, in contrast to standard pairwise transmission dynamics.

We formulated the impact of social reinforcement within a simple spatial SEIRS modeling framework for COVID-19 transmission ^37^. Our COVID-19 transmission model spitted the host population into 4×*n* compartments according to their COVID-19 infection status and their located provinces: with *S_i_*, *E_i_*, *I_i_*, and *R_i_* representing susceptible, exposed, infectious, and recovered portions of individuals in province *i*, where *i*∈{1,…,n} and *n*=31 was the total number of provinces in mainland China. Compartments *S_i_* was further subdivided into two subpopulations by considering individual status for social reinforcement in collective interactions. Specifically, *S_i_*_1_ and *S_i_*_2_ represented susceptible individuals without and with social reinforcement in interactions, where a proportion 𝑝_&_ was used to denote the subpopulation that has been impacted by social reinforcement for province *i*. To assess the impact of heterogeneity in collective interactions on susceptibility, we hypothesized that groups of susceptible individuals engaging collectively in effective contacts with multiple infectors elevated the population risk of infection. This was incorporated by defining the relative FoI for sub-compartments as *S_i_*_2_ , by *ϕ_i_λ*_&,_(𝑡), where *ϕ_i_* denoted the hazard ratio which is modulated by the cluster heterogeneity for province *i* ^38^. To quantify the impacts of social reinforcements on transmissibility, we posited that two infectors under social collective interactions may exhibit augmented transmissibility, and were, therefore *v*(*t*) times more likely to transmit viruses. In addition to capturing collective interactions, our framework incorporates human mobility, viral evolution, and the strength of non-pharmaceutical interventions (NPIs). The model is schematically illustrated in Fig. 1c. Detailed mathematical formulations are provided in the Methods section, with parameter descriptions summarized in Table S1. We used Bayesian inference with the Hamiltonian Monte Carlo (HMC) method ^39^ to calibrate in predicting weekly COVID-19 incidence data in mainland China.

## Epidemiological impact of social reinforcement

To quantify the epidemiological impact of social reinforcement on SARS-CoV-2 transmission, we parameterized the respective impacts (clustering, susceptibility and transmissibility impacts) in the model for predicting the reported incidence dataset (Figs. S1 and S2 in supplementary), thereby capturing the transmission dynamics reflective of spatial characteristics across provinces. Figs. 2a–c presented the proportion of infections attributable to social reinforcement (*p_i_* ), together with the resultant effects on susceptibility (*ϕ_i_* ) across 31 provinces, and the baseline transmissibility impacts (𝑣*_j0_*) for three periods, i.e., from 1 January 2020 to 6 March 2021 for ancestral strains; from 7 March 2021 to 31 December 2021 for Alpha/Delta variants; and from 1 January 2022 to 28 February 2023 for the Omicron variant (Extended Data Fig 1a), respectively. This mechanistic framework is finally able to provide unequivocal insight into the role of social reinforcement in shaping transmission dynamics. Estimates for 𝑝_&_ revealed that 5.3% to 14.4% of individuals in susceptible population were affected by social reinforcement under collective settings. Furthermore, estimates for *ϕ_i_* indicated that social heterogeneity was associated with a multiplicative effect in susceptibility risks ranging from 1.17 (95% CrI: 1.02, 3.66) to 1.71 (1.03, 4.40) times across provinces. Estimates for the variant-specific *v_j0_* showed that transmissibility was augmented by 1.68 (1.03, 2.11) to 1.70 (1.34, 2.24) times comparing with pairwise interaction across the periods. The posterior distributions for the affected proportions and the increased susceptibility across provinces were presented in Extended Data Fig. 2.

**Figure 2.**
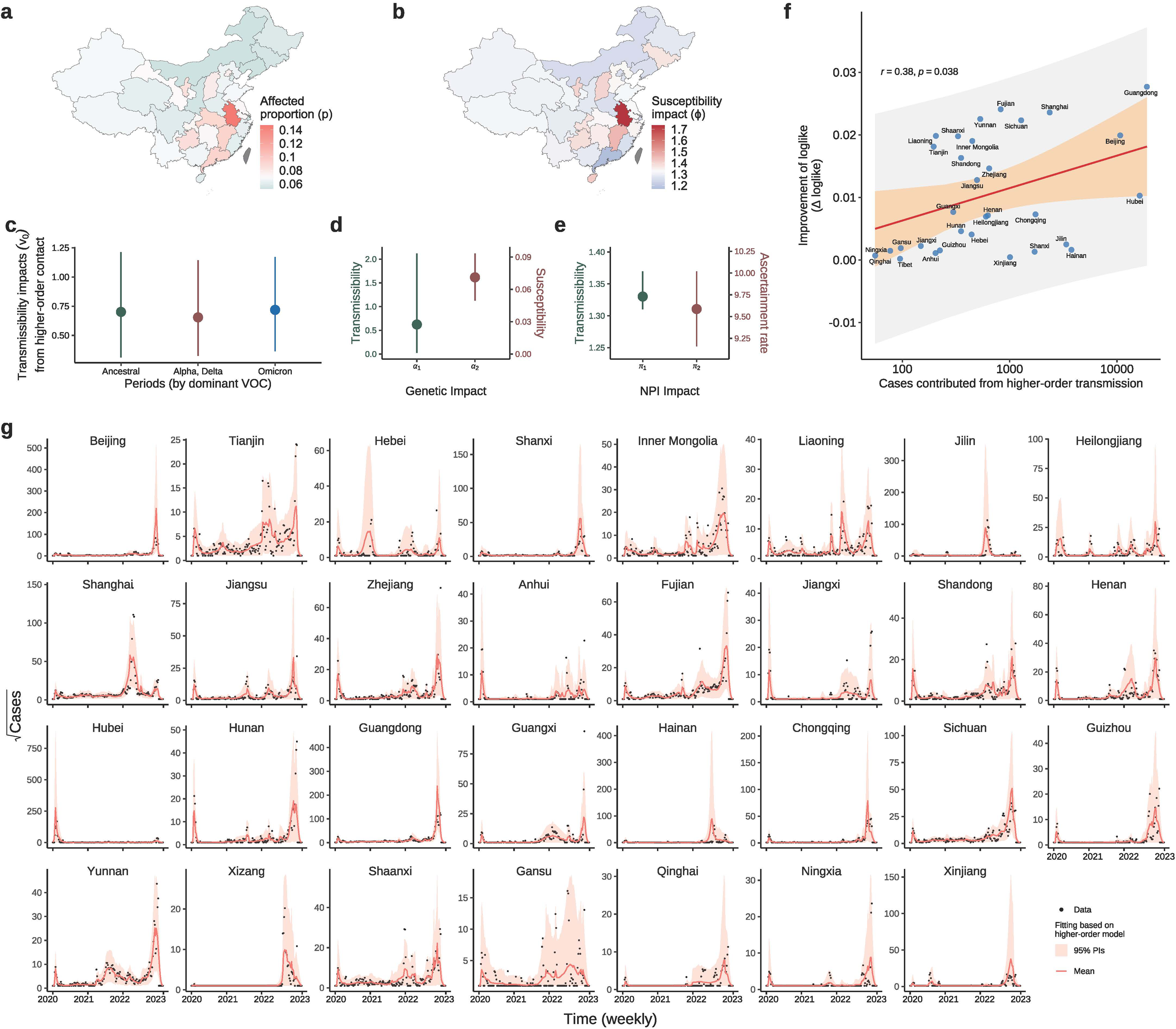
Effects of social reinforcement and higher-order model predictions across 31 provinces during the COVID-19 epidemic. **(a)** Proportion of infection cases influenced by social reinforcement mechanisms across provinces. **(b)** Provincial variation in the impact of cluster heterogeneity on susceptibility. **(c)** Effects of collective interactions on SARS-CoV-2 transmissibility across distinct epidemic periods characterized by the predominance of different SARS-CoV-2 variants of concern. **(d)** The impact of viral genetic diversity on transmissibility and the impact of viral antigenic drift on susceptibility. **(e)** The impact of non-pharmaceutical interventions on transmissibility and ascertainment rates. **(f)** Improvement in predictive performance of the higher-order model relative to the standard model as a function of higher-order transmission, where the red line denoted the predicted mean with 95% confidence intervals (CIs) in orange shade and 95% prediction intervals (PIs) in gray shade. **(g)** Model fit to weekly reported case numbers (black dots) for 31 provinces in mainland China; red solid lines represent mean model estimates and shaded regions indicate 95% PIs.

Moreover, the estimate of coefficients 𝛼_+_ indicated that genetic diversity (Extended Data Fig. 1b) was found to increase the transmissibility under collective interactions by 0.89 (95% CrI: 0.03, 7.76) units per unit change. Our estimate of 𝛼,suggested the antigenic drift (Extended Data Fig. 1c) would increase susceptibility by 0.07 (0.05, 0.10) units per unit change (Fig. 2d). In addition, the estimates of 𝜋_+_ and 𝜋_+_ indicated that per unit increase in NPI (Figs S3-S4) could reduce transmissibility by 2.78% (2.70%, 2.93%) units and increase ascertainment rate by 10.05% (9.58%, 10.52%) units respectively (Fig. 2e).

For model validation, we compared model performance between the higher-order and standard traditional models using identical datasets by assessing improvements in model likelihoods (Δ 𝑙𝑜𝑔𝑙𝑖𝑘𝑒) (Fig. 2f). We observed a significant positive correlation (P-value = 0.007) between the magnitude of model improvement and the number of infections contributed from higher-order infection. Therefore, on average, higher-order model yielded better explanatory power in model-prediction by fitting observed case data (Fig. 2g) for recapitulating the timing and magnitude of distinct epidemic waves at the provincial level.

## Spatiotemporal infection risk transmission

We reconstructed time-varying spatial transmission networks, determined by human mobility and population-level infection risk (details are in Methods section), to quantify the spatiotemporal variation of SARS-CoV-2 transmission, delineated by the distinctive characteristics of infection flows (IFs). As a proxy, we integrated the number of exposed infectious individuals *E_i_*(*t*) and unreported infectious population (1-*ρ_i_*(*t*)) *I_i_*(*t*), where *ρ_i_*(*t*) was the reporting rate for province *i*, mapped onto evolving mobility networks over time *t*. Fig. 3 illustrated the in- and out-bound IFs-based network *eigenvector centrality* during the pandemic at the spatiotemporal scale. In initial stages, Hubei province emerged as the primary *epicenter* of the outbreak for ancestral strain with an estimated total exported IFs of 1343015 (1266504, 1142249), by 6 March 2021 (Fig. 3a). As pandemic progressed, the principal locus of exported IFs shifted to Shanghai during the Alpha/Delta period (March–December 2021), accounting for a cumulative exported IFs of 72756 (61267, 38193). During the subsequent Omicron wave (until February 2023), transmission hubs became increasingly diffuse, with Guangdong emerging as the dominant source, contributing an estimated 5109959 (4924523, 5306426) exported IFs during this large-scale outbreak.

**Figure 3.**
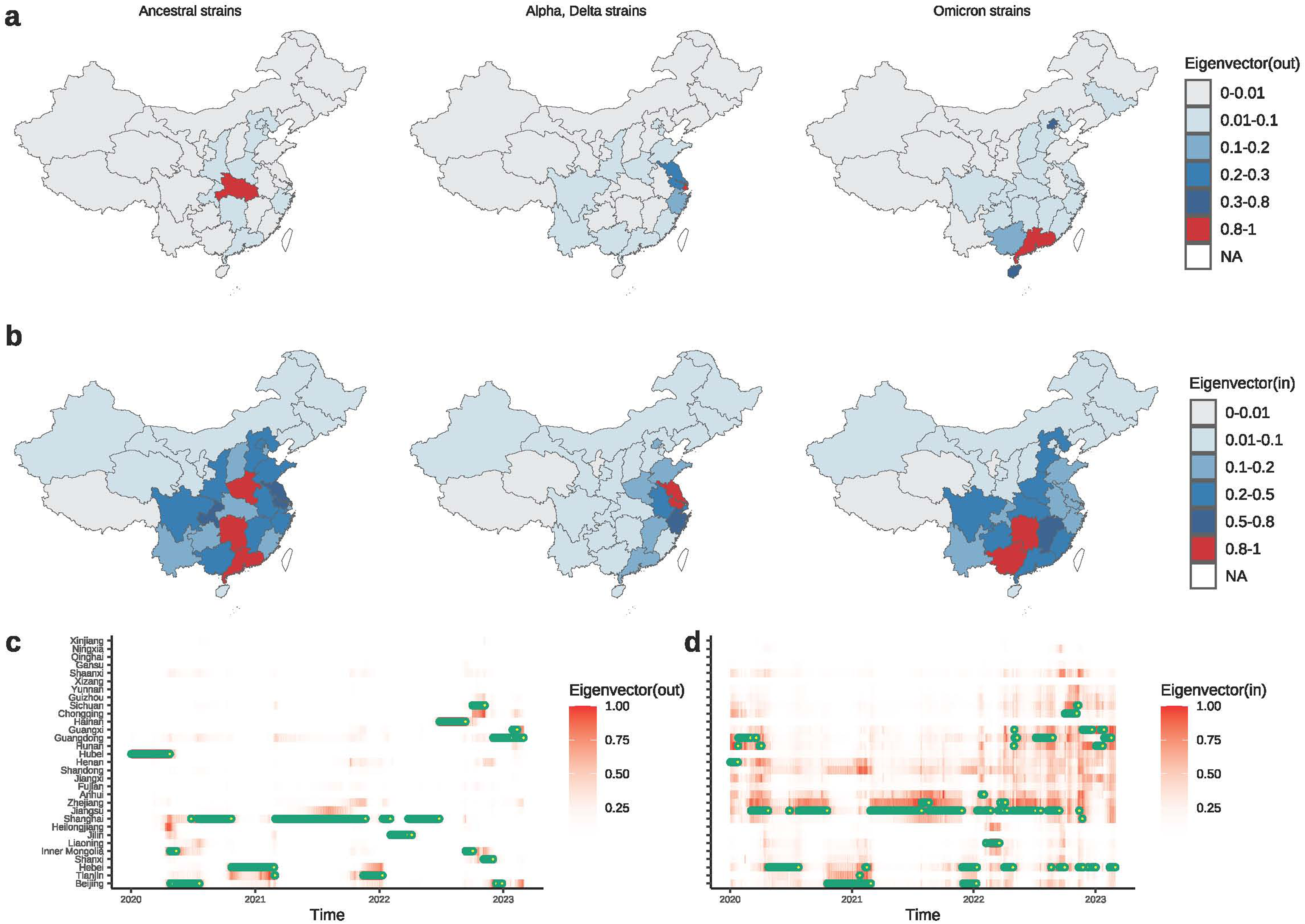
Identification of key transmission locations for SARS-CoV-2 in mainland China, from January 2020–to February 2023. **(a)** Spatial distribution of origin locations, as determined by eigenvector centrality within the outgoing transmission network, highlighting provinces with the highest outflow of SARS-CoV-2 for the three major variants of concern. **(b)** Spatial distribution of key destination locations, based on eigenvector centrality for incoming transmission, indicating provinces with the highest inflow of SARS-CoV-2 associated with the three variants of concern. **(c)** Temporal evolution of dominant source (outflow) provinces identified as vital transmission origins. **(d)** Temporal evolution of major sink (inflow) provinces representing crucial destinations for viral spread. Greater intensity of red in the index corresponds to increasingly significant locations, while blue specifically denotes the most critical transmission hubs at each time point.

Similarly, the earliest principal sink regions were those that absorbed the majority of imported infections: including Henan, Hunan, and Guangdong during initial period with estimated imported IFs of 227464 (213593, 241663), 219950 (208834, 231827), and 175293 (169758, 181992) respectively (Fig. 3b). Subsequently, the importation hubs moved to Jiangsu province during the Alpha/Delta period with an estimated imported IFs of 33932 (28901, 40070). With the onset of the Omicron wave, the scope of importation expanded, encompassing a wider array of provinces; notably, Hunan (1272795 (1226265, 1317602)) and Guangxi (1256078 (1215403, 1299642)) emerged as leading provinces in importing IFs. The spatiotemporal variation of IFs distributions across affected regions at bimonthly intervals were presented in Extended Data Figs. 3–4. The dynamic configurations of source and sink regions, derived from alternative centrality measures such as *degree*, *hub*, and *PageRank centralities*, exhibited comparable characteristics across three periods (Supplementary Figs. S5–S7). In addition, comprehensive visualizations of bimonthly spatial and temporal fluctuations in imported IFs were provided in Figs. S8–S13.

To identify key transmission pathways, we evaluated *link weight centrality* and *betweenness centrality* over time, enabling us to track the temporal evolution of network connectivity across three pandemic periods (Fig. 4) as well as at bimonthly intervals (Extended Data Figs. 5-6). During the ancestral strain period, the principal transmission pathways, quantified by *link weight* found to be managed from Hubei, extending predominantly to Henan (exported IFs: 209392 (195918, 222980)) and Hunan (exported IFs: 209528 (198511, 220923)) (Fig. 4a).

**Figure 4.**
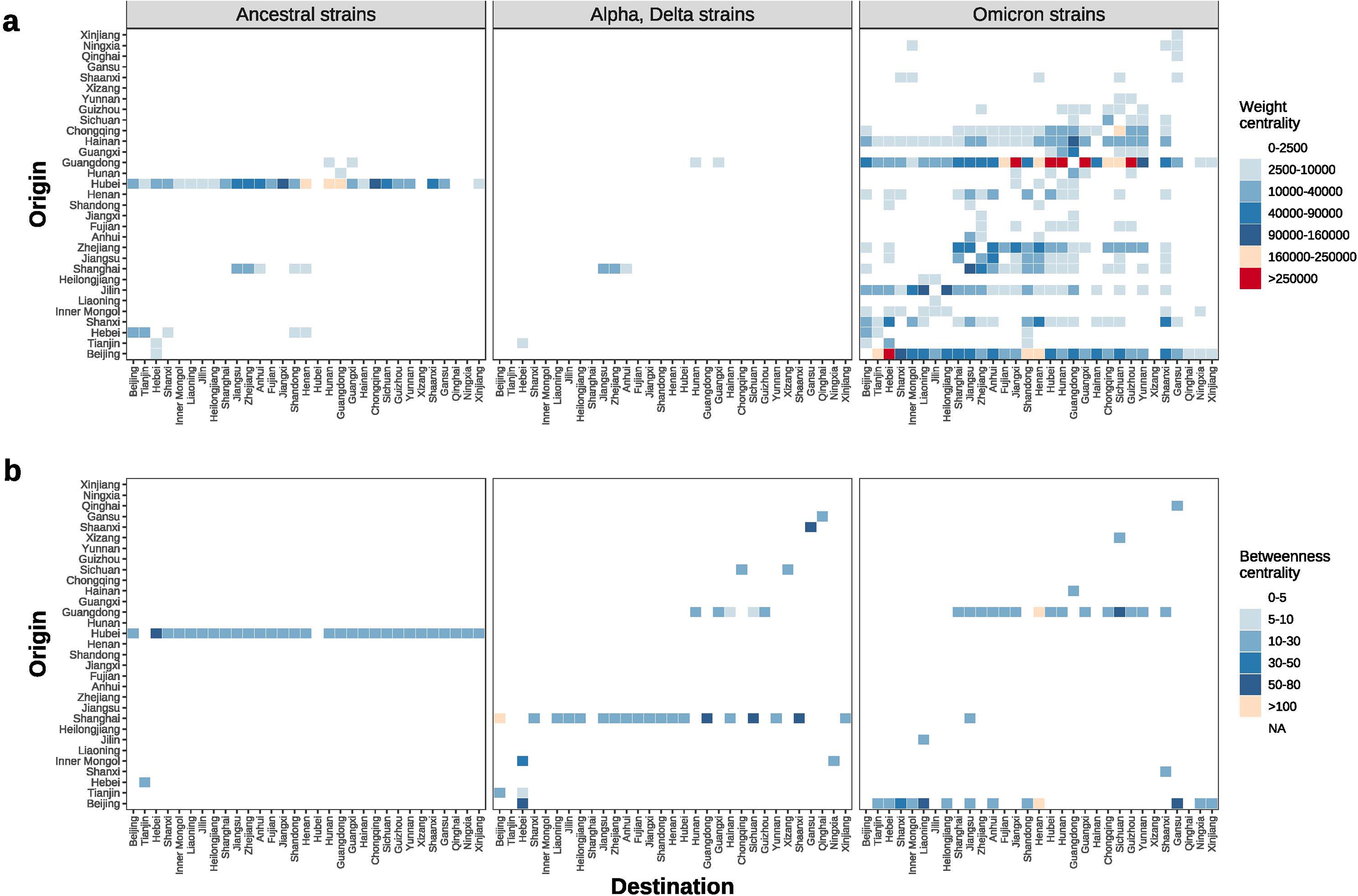
Key link characteristics of SARS-CoV-2 transmission networks across three epidemic periods in mainland China, from January 2020 to February 2023. Link importance within the transmission networks is quantified using link weight centrality **(a)** and betweenness centrality **(b)** for three distinct epidemic periods. Higher values denote greater structural and epidemiological importance of transmission pathways.

These dominant pathways underwent a marked shift during the Alpha/Delta period, with the primary route connecting Shanghai and Jiangsu and a corresponding exported IFs 30288 (25510, 36182), substantially lower than that observed in the ancestral period. During Omicron period, transmission networks grew considerably more complex and spatially diffuse, giving rise to multiple provincially interlinked hubs as key locations of viral dissemination. Notably, the IFs from Guangdong to Fujian (exported IFs: 562761 (540650, 586860)), Hunan (1061837 (1015522, 1111964)), and Guangxi (1179367 (1137934, 1224628)) constituted major pathway of infections, significantly higher of those risks estimated during earlier periods, attributing the impact of the lifting of the “zero-COVID” policy in December 2022. Similarly, using network *betweenness centrality*, defined by the frequency with which a link appeared along shortest paths between all pairs of locations, we identified the Hubei–Hebei connection as particularly indispensable during the ancestral period (Fig. 4b). In the Alpha/Delta period, the central bridge pathway shifted to connect Shanghai and Beijing. The Omicron surge was characterized by further complexity and comparatively wide dispersal, with the most critical pathways emanating from Guangdong and Beijing toward Hainan.

## Time-varying geographic cluster distributions and the relationship between infection outcomes and human mobility

Spatiotemporal analysis of inter-provincial transmission networks demonstrated that the geographic cluster distributions varied across three pandemic periods (Figs. 5a, 5e, 5i). Variation in geographic cluster distributions contributed to differences in infection risk and associated transmission across provinces. Notably, provinces within the same cluster were often geographically adjacent, highlighting the role of land-based transportation in shaping transmission dynamics. Although geographic cluster structure changed over time, the main clusters of infection risk flow consistently corresponded to the populous regions: northern, southern, and eastern provinces of China.

**Figure 5.**
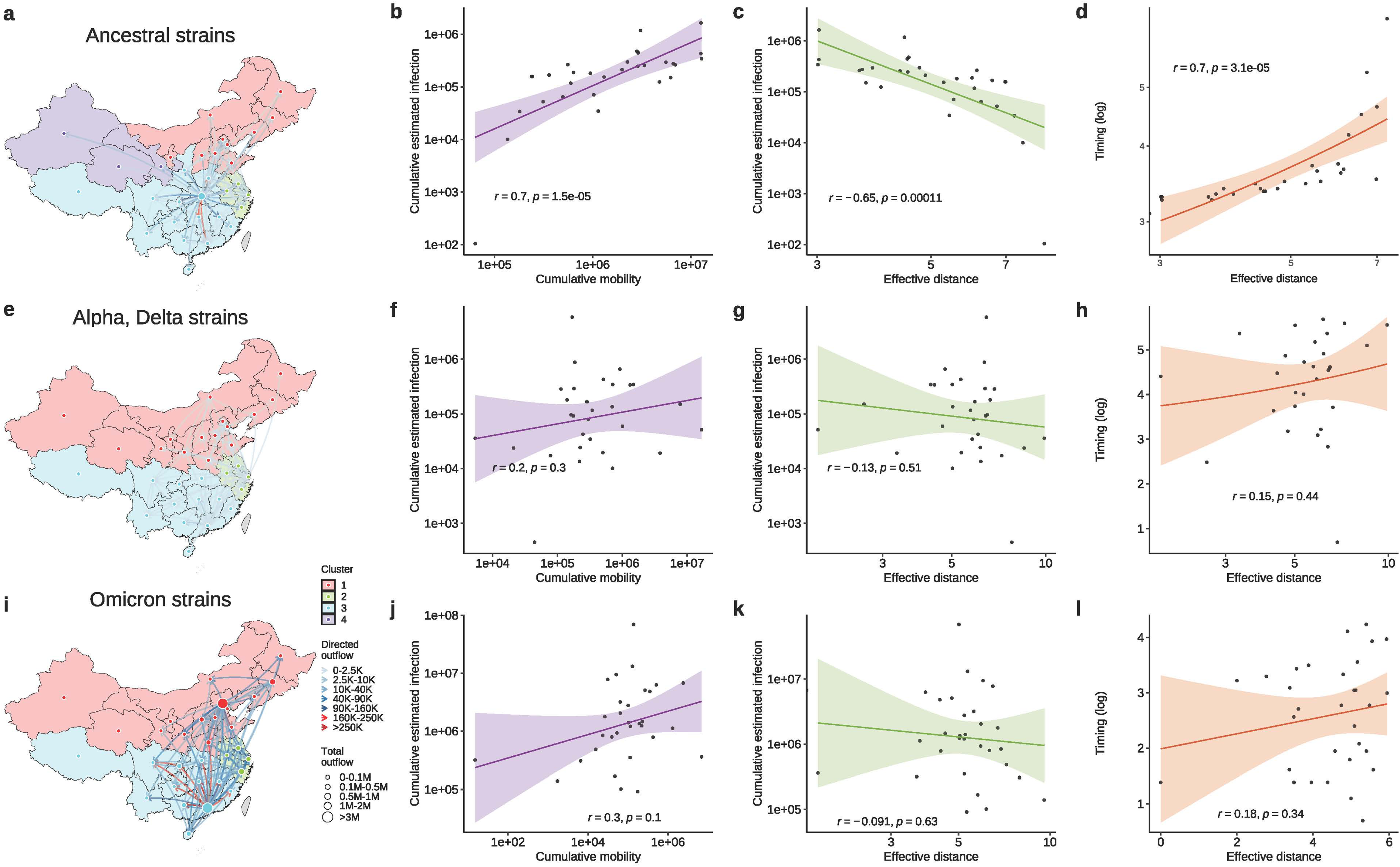
Time varying geographic clusters of transmission networks and associations between human mobility and SARS-CoV-2 transmission dynamics across three pandemic periods in mainland China, from January 2020 to February 2023. **(a)** Geographic clustering of provinces within the transmission network during the ancestral strain period; provinces sharing the same color belong to the same cluster, and node size reflects the magnitude of infection risk outflow. **(b–d)** Relationships during the ancestral strain period: **(b)** reported cases versus inter-provincial mobility, **(c)** reported cases versus effective distance, and **(d)** time to reach 100 cases versus effective distance in the early epidemic period. **(e–h)** and **(i–l)** present analogous analyses for the Alpha/Delta and Omicron variant periods, respectively, illustrating changes in cluster structure and associations between mobility patterns and transmission characteristics for each period.

During the ancestral strain period, human mobility patterns associated with IFs centered on Hubei, which functioned as the primary source hub, with node and link sizes proportional to mobility volume (Fig. 5a). In the Alpha/Delta period, Shanghai emerged as a significant hub, though its influence and overall mobility volume were reduced (Fig. 5e). By the Omicron period, hub shifted to multiple provinces, notably Guangdong and Beijing, both exhibiting markedly increased IFs volumes (Fig. 5i). At bimonthly temporal resolution, an intriguing pattern emerged whereby, during the winter seasons (January–February) except 2021, Hainan Province in southeastern China clustered with provinces in Northeast China, likely reflecting seasonal migration as populations from the Northeast moved to Hainan to escape winter conditions (Extended Data Fig. 7).

Furthermore, we quantified the association between IFs outcomes (magnitudinal and temporal) and human mobility under the predicted transmission networks, including the cumulative number of infection cases (final size) and the timing required to reach specified case thresholds across the periods (Fig. 5). During the ancestral period, Hubei acted as the main transmission hub (Fig. 3a). To capture the mobility patterns related to this hub, we assessed how the number of travelers from Hubei was linked to reported COVID-19 cases in other provinces. The predicted provincial cumulative case counts strongly correlated (*r* = 0.7, P-value < 0.005) with the volume of inbound travelers from Hubei (Fig. 5b). We evaluated *effective distance* ^40^, which quantifies inter-regional connectivity based on travel flows, to better capture how transmission relates to population mobility. The estimated final size for the period of ancestral strains found to be negatively associated (*r* = -0.65, P-value < 0.005) with effective distance from Hubei Province (Fig. 5c), while a positive association (*r* = 0.7, P-value < 0.005) was found between the time to reach first 100 predicted cases following the initial outbreak in Hubei (1^st^ January, 2020) and their effective distance (Fig. 5d). Comparable results found on the relationships for observed final size on mobility and effective distance for ancestral period in China (Extended Data Fig. 8). However, during Alpha/Delta and Omicron waves, these associations found to be weaker (but same directions) and insignificant (Figs. 5f-h and 5j-l), suggesting the role of other factors confounded as pandemic progressed.

## Discussion

The higher-order modeling framework could assess and quantify the potential role of collective interactions within clustered social settings in shaping epidemic trajectories, assimilating social reinforcement under a simplicial complex ^13,24,27^. In this framework, several settings, such as workplaces, restaurants, and households are included as higher-dimensional units (simplices) to account for the impact of transmission heterogeneity ^9,24^. This formulation allowed for the explicit representation of collective interactions, rather than only pairwise interactions, and overcame the limitation of the common linear assumption of infectious contacts on infection risks. By capturing the higher-order topology of social contacts, typically overlooked in standard models, our framework enables rigorous identification of potential social determinants of epidemic transmission that are inaccessible to standard approaches (Fig. 1a-b) ^9,10,24^. When coupled with empirical datasets on genetic diversity, population mobility, our model substantially enhances the granularity at which spatiotemporal transmission dynamics can be resolved at various scales (provincial, national, and global) (Fig. 1c) ^17,23,26–29^. Crucially, the proposed higher-order modeling framework outperformed standard models in predicting pandemic trajectories ^24^, offering demonstrably superior explanatory and predictive power (Fig. 2f).

Social reinforcement revealed pronounced heterogeneity in both the proportion of infections attributable to such reinforcement and the resultant increases in susceptibility across provinces (Fig. 2a-b). Notably, while the proportion of infections influenced by social reinforcement generally remains modest, ranging between 5.3% and 14.4% across mainland China, its impact can be substantial, serving as a catalyst for critical phenomena such as superspreading events ^6–8,13,24,35^. Provinces exhibiting pronounced clustering manifested increased susceptibility up to 17% to 71% higher risk than those observed under pairwise mixing regimes. Within these clustered settings ^41^, susceptibility demonstrably escalates with increasing population size, a trend robustly captured by our higher-order modelling framework, which partitions the susceptible population into discrete subgroups, thereby extending beyond the classical assumption of pairwise interaction ^9,10,13,24,29^. These findings contribute to a deeper understanding of the underlying mechanisms driving clustering and superspreading phenomena ^9,29,42^.

Furthermore, the higher-order contacts found to consistently escalate the transmissibility of COVID-19 and consistent across the periods (Fig. 2c). One possible reason is that collective interactions, which involve exposure to a greater number of infectious individuals, may result in a higher cumulative dose of viral particles, surpassing the capacity of the human immune system ^9,13,29,43^, thereby concurrently increasing the transmissibility. Crucially, transmissibility driven by social reinforcement increases by more than half (68%–70%) compared to transmission seen in pairwise contacts. Although it is rare for several infected people to be in the same group when the overall infection rate is low, such events can spark major outbreaks and superspreading ^9,29,42^. These findings support the significance of non-pharmaceutical interventions, such as social distancing and restrictions on large gatherings, in controlling the spread of respiratory infectious diseases ^30,32^.

A nationally representative, population-based examination of the spatiotemporal variation and unique characteristics of SARS-CoV-2 transmission would significantly deepen our understanding of epidemic dynamics on both temporal and geographic scales ^2^. By reconstructing transmission networks derived from the spatial distribution of infectious population and population mobility at temporal scale, our analysis reveals spatiotemporal epidemiological characteristics, including critical locations (Fig. 3), key patterns of infection flows (Fig. 4), and underlying geographic cluster structures (Fig. 5) across the provinces in mainland China. We could identify several underline significant factors for the spatiotemporal transmission pattern of COVID-19, which vary markedly across the provinces during three predefined pandemic periods. In particular, the population mobility from epidemiological hubs found to increase the infection rates in downstream (sink) regions by elevating the risk of secondary infection (Fig. 5). The provinces with higher traveler volumes and shorter effective distances from hub locations experienced significantly greater infection risks and earlier epidemic onset. However, the strength of these associations declined during the Alpha/Delta (Fig. 5f–h) and Omicron (Fig. 5j–l) periods, reflecting a diminished influence of centralized transmission hubs ^40,41,44^. This temporal shift indicates a gradual decentralization of transmission pathways as the epidemic progressed, marked by the emergence of multiple, simultaneous outbreaks. Unveiling principal locations, pathways, geographic clusters, and the relationship between infection and population mobility would help us achieve effective and targeted control of epidemic transmission in large countries and at the global level ^2,4,45^.

Despite within-host viral interactions, competition, and mutation, the presence of multiple infectors in a group, each carrying viruses with distinct genetic backgrounds, amplifies genetic diversity and facilitates inter-viral interactions ^23^. These interactions promote viral competition and mutation, collectively enhancing the potential for transmission dynamics by reshaping the associated susceptibility and hence transmissibility ^17,23^. Our findings demonstrate that greater genetic diversity is associated with increased transmissibility under collective interactions (Fig. 2d). Furthermore, antigenic drift alters population immunity, a property we term immune efficacy with respect to susceptibility ^46,47^, and is associated with increased susceptibility as ongoing antigenic changes diminish the effectiveness of host immune responses (Fig. 2e).

The impact of NPIs stringency is evident in the attenuation of transmission rates, as demonstrated in Extended Data Figs. 9-10. We found that the increase in NPI stringency is associated with a reduction in the transmission rate, with the corresponding effects on the effective reproductive number, comparable to previous reports ^32,33^, illustrated in Figs. S14–S15. Besides, NPI stringency is positively correlated with improvements in case ascertainment, as shown in Figs. S16–S17. Collectively, these results highlight the dual function of NPIs in both mitigating viral transmission and enhancing the rates of infection detection ^48^.

Our higher-order modelling framework that integrates viral genomic data, non-pharmaceutical interventions, and human mobility to quantify the effects of collective interactions and spatiotemporal heterogeneity within subpopulations, yields demonstrably improved predictive performance over conventional epidemic models. The flexibility of this paradigm highlights its broad applicability across diverse geographic settings and to other respiratory pathogens. Our study has limitations. First, we identify population mobility as a principal driver of the observed spatiotemporal variability in epidemiological dynamics. Although the use of mobility data from Baidu Qianxi substantially enhance inference ^33^, fully capturing the complexity of real-time, nationwide inter-regional mobility remains challenging. Second, while vaccination coverage is high, but its impact on disease infection rates appears to be mild to modest ^49^ and vaccination schemes were centralized with minimal variation in timing and coverage across the provinces in mainland China ^50^. To focus on our primary objectives, we therefore opted out the vaccination impact in the model directly ^2,18^, however availability of such data could be tested for the model performance. Third, the limited availability of fine-grained cluster data precludes robust characterization of transmission dynamics within specific settings, such as workplaces or restaurants, across China at temporal scale. Therefore, agent-based analyses could be limited in such case; however, the individual-level investigations could be a better alternative^24,51^. Fourth, while our simplicial models with up to order 2 interactions offer an important insight into the hidden mechanisms of infectious disease transmission. However, considering higher-order interactions (over order 3 or beyond), could be a pivotal to capture transmission dynamics better. Future research could account the contribution of such interactions of varying orders for transmission heterogeneity ^9,10,24^. Finally, we considered the nationwide mobility datasets in mainland China ^52^, nevertheless the inclusion of international mobility data could provide better importation risk prediction over time. Although, international travel restrictions might reduce such risk of infection during the study period.

In conclusion, we could establish a data-driven, higher-order modelling framework that assimilates epidemiological, geographic, genomic data steams with complex network approaches to delineate the foundational role of social reinforcement in shaping collective transmission dynamics and the spatiotemporal evolution of transmission networks across regions and epidemic phases. Understanding the impacts of social reinforcement at various levels of transmission process and other potential factors would provide a perspective tool for mitigating the burden of COVID-19 and other respiratory pathogens and their emerging variants under heterogeneous population settings.

## Methods

### Higher-order modelling with social reinforcements for collective interaction

Inspired by a simplicial higher-order model ^9,10,24^, we accounted for both standard homogeneous interactions and heterogeneous clustered group interactions to identify the impact of social reinforcements. We constructed the following deterministic skeleton of the model, incorporating collective interactions for social reinforcement and demographic profiles (Fig. 1). The spatiotemporal dynamics of the transmission process are subsequently described within this deterministic modeling framework.

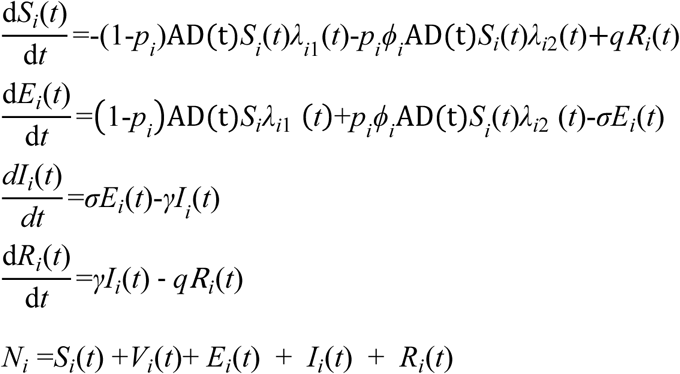

Where *S_i_*(*t*),*E_i_*(*t*),*I_i_*(*t*) and *R_i_*(*t*) denoted the proportion of susceptible individuals, exposed individuals, infectious individuals, and recovered individuals at time *t* for province *i* , respectively. AD(t) = 𝑒^𝛼^_2_ ^*ad(t)^ denoted the impact of antigenic drift on susceptibility, where ad(t) was the time-varying antigenic drift index and 𝛼, denoted its impact. An infected individual would become infectious after 1/*σ* days on average since infection (i.e., mean latent period), which was assumed to be 5 days ^2,24,53^. The individual would be infectious for 1/*γ* days on average (i.e., mean infectious period), which was assumed to be 10 days ^2,24,53^. The duration of sterilizing immunity 1/*q* was assumed to be 610 days ^46^. *p_i_* represents the proportion of infections influenced by social reinforcement under the higher-order collective interactions for province *i* and *ϕ_i_* denotes the impact of social reinforcements on susceptibility. Note that when *p_i_*=0, the high-order model will be degenerated into the standard homogeneous model. The force of infection in standard and higher-order transmission settings with IFs can be defined as follows:

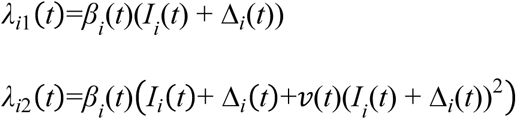

where *β_i_*(*t*) was the time-varying transmission rate for province *i*. *v(t)=*𝑣*_j_*_0_ (𝑡)𝑒^𝛼1\**div*(*t*)^ denoted the transmissibility impact under social reinforcements where more than one infector are in the same cluster, and time *t* is segmented by the timing of the leading variants of concern. Here, 𝑣_!"_(𝑡) represents the baseline impact of social reinforcement for dominant variant of concern 𝑗 and div(𝑡) denoted the genetic diversity data and 𝛼_+_ was its impacts. The impact of human mobility on provincial infectiousness is quantified by combining mobility-driven transmission dynamics with spatiotemporal network properties, computed as follows, 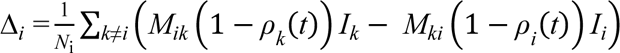,where *M* is the population flow from province *k* to province *i*, and *ρ_i_*(*t*) denotes the ascertainment rates (i.e., reporting proportion) of province *i*. Here, we assumed that mobility was restricted for documented infections.

We constructed the time-varying transmission rate *β_i_*(*t*) for province *i* at time *t* as a function of cubic *B-spline* and non-pharmaceutical interventions (NPIs), as *β_i_*(*t*)= *exp*(*s_i_*_,*t*_) * *β_NPI_*_, *i*_(*t*), where *s_i_*_,*t*_ is a degree 3 spline function with 10 equally spaced knots based on the whole study period for the province *i* at time *t*. *β* (*t*)=𝑒^-π^_1_*^NPI(i,t)^* denotes the impact of the NPI and 𝜋_+_ represents the level of impact and *NPI(i*,*t)* represents the NPI stringency index for province *i* at time *t* . We assumed the time-varying ascertainment rates (i.e., reporting proportion) *ρ_i_* for province *i* is often driven by the associated NPIs ^54,55^. Therefore, assuming a positive association between the strength of NPI and ascertainment rates, we used a sigmoid function to describe time-varying ascertainment rates as *ρ*_i_ (*t*)*= ρ* 𝑒^-π^_2_*^NPI(i,t)^* where 𝜋, denotes the impact level and *ρ*_0_ denotes the baseline ascertainment rate without the impact of NPI (i.e., *NPI(i*,*t)*=0) in the settings.

### Model inference framework

To relate the predicted incidence to the empirically observed COVID-19 cases, let *C_i_*(*t*) be the cumulative observed cases at week *t* for province *i*, defined as 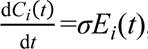, accordingly, the observed cases 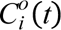 at time *t* for location *i* was derived as 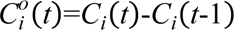. Further, we described the observed incidence *y_i_*(*w*) at week *w* as a negative binomial process of the model induced cases as: 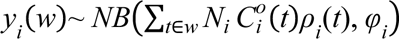, where 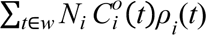 denotes the mean of predicted cases at week *w*. *N_i_* is the total population of province *i* and *φ_i_* is a parameter denoting overdispersion.

Model fitting and estimation of parameters *θ*={*s_i_, p_i_, ϕ_i_, v(t), ρ_0_, a, b, c, d, φ_i_*} were conducted within a Bayesian inference with Hamiltonian Monte Carlo (HMC) ^39^. In this approach, prior distributions for certain model parameters were specified conditionally, reflecting the nature of the Bayesian paradigm. Uninformative priors were incorporated throughout the inference process (Supplementary 2). We employed the *RStan* package in R, leveraging the No-U-Turn Sampler (NUTS) algorithm ^39^. Four independent Markov chains were initiated at different staring positions, each comprising 4,000 iterations with the first 2,000 iterations designated as burn-in. Convergence and thorough exploration of the posterior distribution were assessed by ensuring that the *R-hat* statistic for all parameters remained below 1.05, thereby indicating satisfactory mixing and convergence ^56^.

### Reconstruction of the time-varying transmission network

In addition to characterizing the general population, we specifically investigated the mobility network of individuals at potential risk of infection. To reconstruct time-varying, virus-carrier-based transmission mobility networks, we integrated data on province-level human mobility with model-inferred, province-specific viral activity. The mobility of population at time *t* between provinces is denoted by a mobility matrix *M*(*t*) with entries *M_ji_*(*t*) representing the number of individuals moving from province *i* to province *j* at time *t* . The infection dynamics in each province are captured by a potential IF matrix, represented as *E_i_*(*t*) + (1 - *ρ_i_*(*t*))*I_i_*(*t*), *E_i_*(*t*) represents the inferred exposed population, assumed to be unrestricted in their mobility, *I_i_*(*t*) denotes the inferred infectious population at time *t*, where the mobility of such reported cases was prohibited, and thus we just considered the unreported infection cases, and *ρ_i_*(*t*) denotes the vector of ascertainment rates for all provinces at time *t*. The transmission network at time *t* can be computed as *T*(*t*) = *M*(*t*) · diag(*E*(*t*) + (1 - *ρ*(*t*))*I*(*t*)), where diag is used to denote a diagonal matrix, 𝐸(𝑡) = {*E_i_*(*t*)}*_i_*_=1…31_ , 𝜌(𝑡) = {𝜌*_i_*(*t*)}*_i_*_=1…31_ and *I(t)*=*{I_i_*(*t*) *}_i_*_=1…31_ . This integrative framework allows for the coupling of inter-provincial mobility patterns with local IF profiles, facilitating the characterization of spatiotemporal virus transmission across provinces.

### Transmission network assessment and spatiotemporal characteristics

The properties of complex transmission networks were primarily characterized by (1) the identification of potential infection hubs—critical nodes or provinces implicated in disease transmission ^33,45,57^; (2) the delineation of high-risk transmission pathways, key mobility links facilitating disease dissemination; and (3) the recognition of transmission geographic clusters—groups of provinces tightly interconnected through frequent exchanges ^58^. The elucidation of these structural elements within transmission networks is essential for informing the design of targeted and effective intervention strategies. To identify putative infection hubs or key nodes, we employed several centrality metrics: (a) *Degree centrality*, quantifying the number of direct connections per node, where provinces with high degree centrality are positioned to facilitate rapid disease spread owing to numerous immediate contacts; (b) *Hub centrality*, as defined by Kleinberg’s algorithm and calculated by *hub score*, providing a measure of a node’s prominence as a key transmitter within the network, reflecting its overall influence on epidemic dynamics; (c) *Eigenvector centrality*, which captures not only the direct connectivity of a node but also considers the influence exerted by its neighbors—provinces with elevated eigenvector centrality are thus both extensively connected and linked to other influential provinces; and (d) *PageRank centrality*, ranking nodes based on both the quantity and quality of incoming connections, thereby highlighting those provinces of greatest strategic importance within the transmission network ^45^. Considering vital locations, we assess key nodes in the transmission network from two perspectives: (1) sources, defined as principal outflow provinces from which SARS-CoV-2 disseminates outward, and (2) sinks, or primary inflow provinces to which infection is introduced.

A suite of well-established centrality metrics was employed to elucidate key links within the transmission network across three distinct periods: (a) *Weight centrality*, which quantifies the strength of interconnections in a weighted network, wherein link weights correspond to the probability or intensity of disease spread—heavier links thereby reflect a greater potential for pathogen transmission ^58^. (b) *Link betweenness centrality*, which assesses the frequency with which a given link appears on the shortest paths between node pairs. Links with high betweenness centrality serve as critical “bridges” within the network, whose removal is likely to fragment connectivity and substantially impede the progression of outbreaks—such links thus represent pivotal routes for transmission ^59^.

To further delineate the structure of transmission, we implemented the walk trap algorithm to detect geographic transmission clusters in time-varying, weighted, directed networks by interrogating node and link interrelationships alongside their contributions to epidemic trajectories ^60,61^. Such clusters typically emerge when groups of infected individuals in one province establish strong connections with those in other provinces, revealing localized outbreaks that share common infection sources. These clusters may function as amplification zones that not only sustain intra-regional spread but also potentiate the exportation of infections to new geographic areas.

### The impact of human mobility on transmission dynamics

To assess the influence of human mobility on the transmission dynamics of SARS-CoV-2, we examined the relationship between human mobility patterns and key epidemiological characteristics of SARS-CoV-2 ^32,40^, including the relationship between cumulative human mobility from the epidemic hub and the number of reported cases, considering both accumulative migration metrics and effective distance cumulative number of infection cases and the timing required to reach 100 reported case thresholds ^40^. Here, the effective distance from province *j* to *i* is defined as ^40^ *d_ij_*=1-log(*P_ij_*), where *P_ij_* is the fraction of flux leaving province *j* that is arriving at province *i*, and therefore can also be written as *P_ij_*= *M_ij_*⁄*M_j_*, where *M_j_*= ∑*_m_ M_mj_* is the total number of population leaving province *j*.

### Ethics statement

This study was conducted in strict accordance with all relevant ethical regulations and guidelines. Ethical approval was obtained from the Institutional Review Board of The University of Hong Kong/Hospital Authority Hong Kong West Cluster (HKU/HA HKW IRB; reference number: UW 22-761). All methods were performed according to relevant guidelines and regulations. Informed consent was obtained from all participants and/or their legal guardians, as appropriate.

### Data availability

We obtained province-level, daily reported COVID-19 surveillance data for mainland China spanning 1 January 2020 to 28 February 2023 from the Johns Hopkins University Coronavirus Resource Center (JHU CRC) (https://github.com/CSSEGISandData/COVID-19) ^2,62,63^. Human mobility patterns over the study period were characterized using data provided by Baidu Corporation (https://qianxi.baidu.com), which represents one of the most widely utilized open-access nationwide mobility datasets in mainland China ^52^. This dataset comprises both the migration scale index and the migration proportion for each province across time. Data on the stringency of non-pharmaceutical interventions (NPIs) were sourced from the Oxford COVID-19 Government Response Tracker (OxCGRT) (https://github.com/OxCGRT/covid-policy-tracker) ^64^, and the stringency index was rescaled to the unit interval [0, 1] for incorporation into the model. Information on SARS-CoV-2 lineages circulating in mainland China during the study period was obtained from publicly available submissions to GISAID (https://gisaid.org/). All genome sequences and associated metadata in this dataset are published in GISAID’s EpiCoV database. Detailed information for each sequence, including accession number, virus name, collection date, originating and submitting laboratories, and authors, can be found under EPI_SET_250805xc. This dataset includes 27,605 genome sequences collected between 2020-01-01 and 2023-02-28. These datasets were utilized to ascertain the temporal emergence in predominance of variants of concern genetic diversity and antigenic drift. Provincial population sizes for 2021 were obtained from the China City Statistical Yearbook (http://olap.epsnet.com.cn/). The geographic location of each provincial unit was determined by the latitude and longitude coordinates of its central point.

## Code availability

All data and code for the study analysis and simulations in this paper will be publicly available from https://github.com/WangDongHKU/high_order_modelling.

## Acknowledgments

The authors thank Julie Au for technical assistance. We acknowledge the research computing facilities and advisory services (HPC) offered by Information Technology Services, The University of Hong Kong and Laboratory of Data Discovery for Health (D^2^4H).

## Author contributions

All authors meet the ICMJE criteria for authorship. Conceptualization: S.T.A., D.W. and B.J.C.; Data curation: Y.W.,Y.T., D.W. and S.T.A.; Methodology: D.W. and S.T.A.; Formal analysis: D.W. and S.T.A.; Investigation: D.W. and S.T.A.; Visualization: D.W. and S.T.A.; Software: D.W.; Validation: D.W., O.G. and S.T.A.; Resources: S.T.A. and B.J.C.; Funding acquisition: D.W., S.T.A. and B.J.C.; Supervision: S.T.A.; Validation: D.W., and S.T.A.; Writing-original draft: D.W. and S.T.A. ; Writing-review and editing: Y.W., J.C., Y.T., H. W., D.C., O.G., E.H.Y.L, Y.Z., P.W. Q.Z. and B.J.C. All authors discussed and approved the manuscript.

---

**Extended Data Fig. 1.**
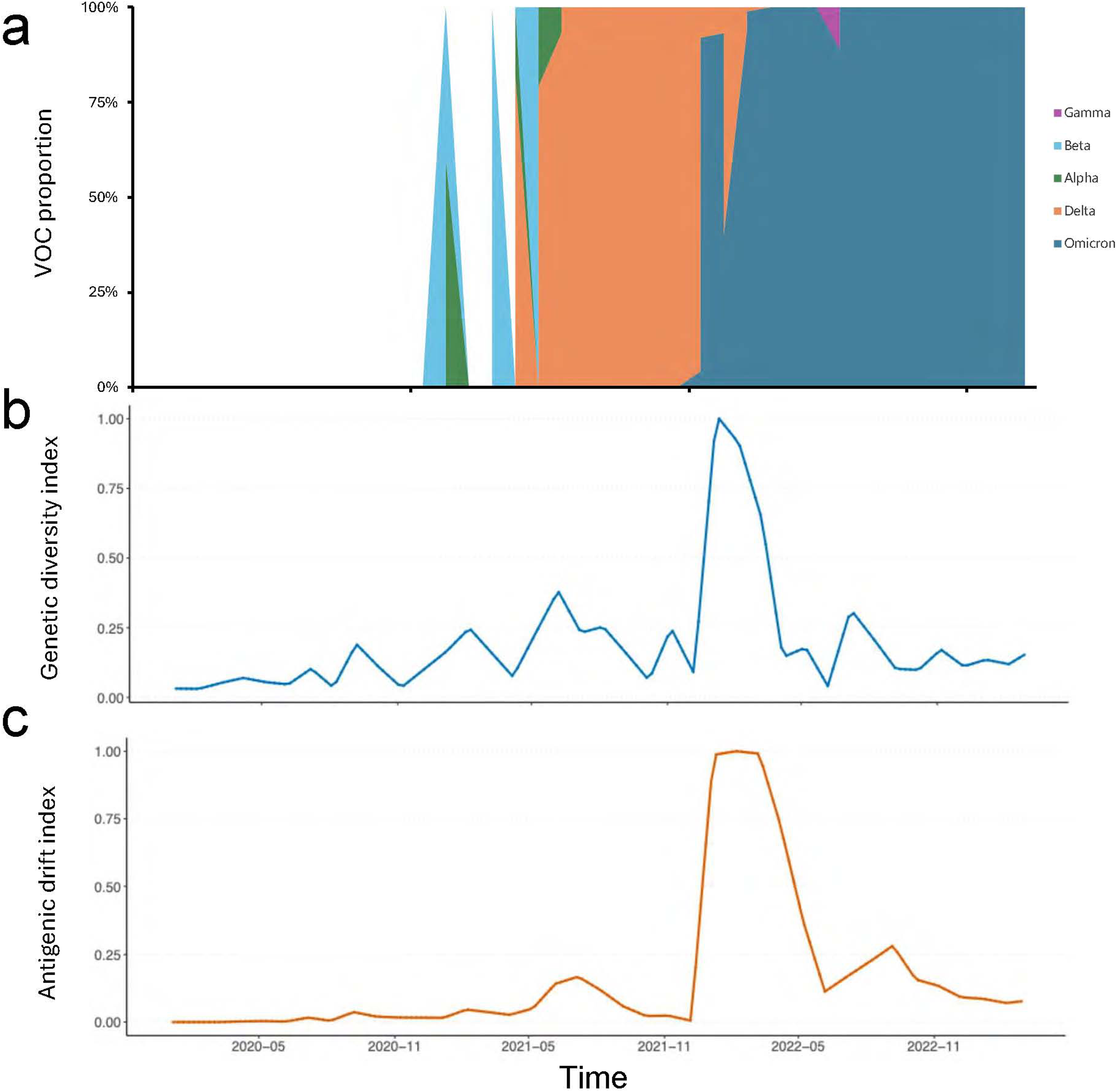
The virus genetic drift data for SARS-CoV-2 from December 2019 to February 2023. **(a)** the proportion of variants of concern, **(b)** genetic viral diversity and **(c)** antigenic drift of SARS-CoV-2 over time. The data sets are collected from GISAID (https://gisaid.org). Originating Lab and Submitting Lab and the list of Authors, visit https://doi.org/10.55876/gis8.250805xc.

**Extended Data Fig. 2.**
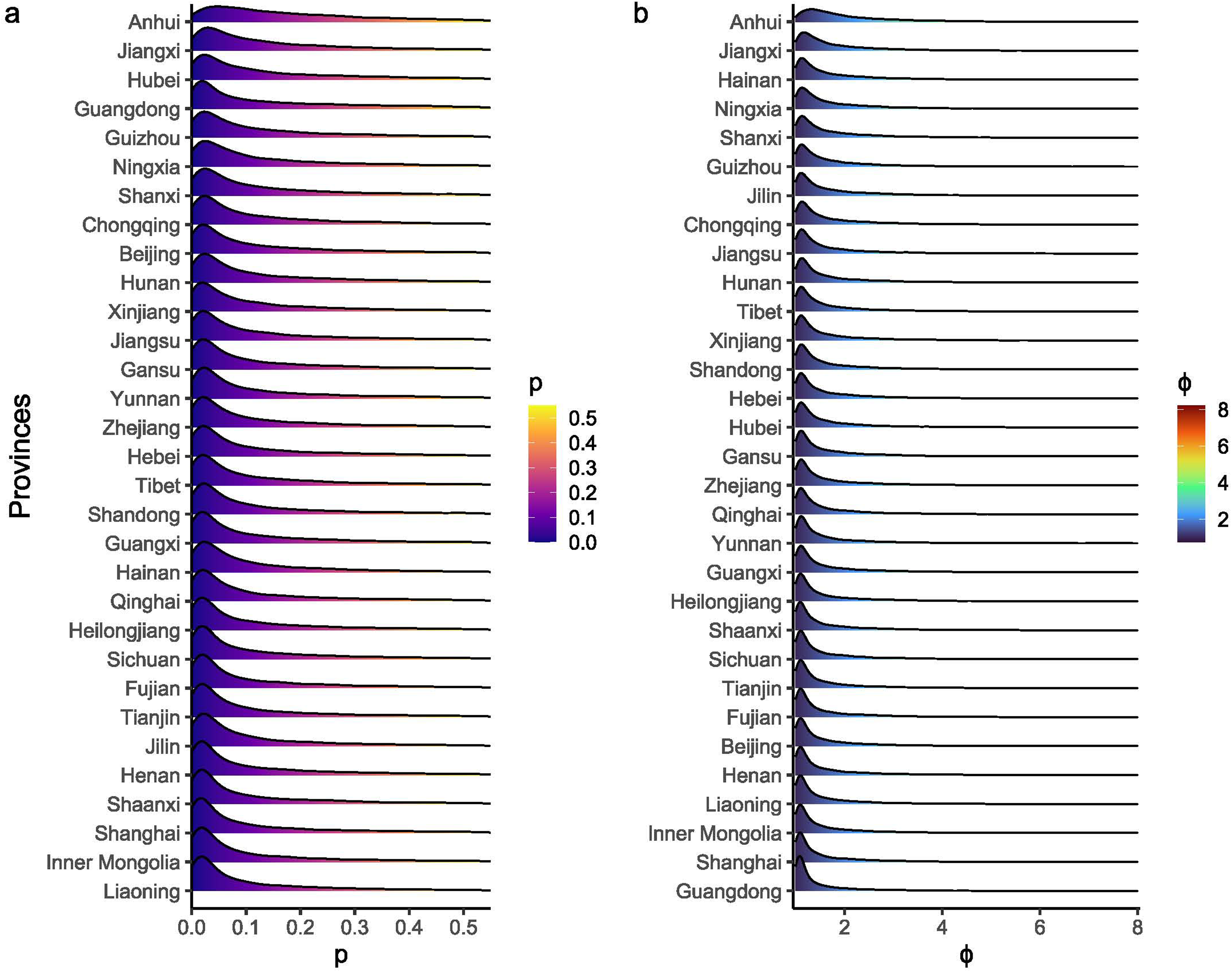
The posterior distributions of transmission parameters across provinces: **(a)** the proportion of infection cases influenced by social reinforcement mechanisms across provinces, **(b)** the impact of collective interaction on susceptibility across provinces.

**Extended Data Fig. 3.**
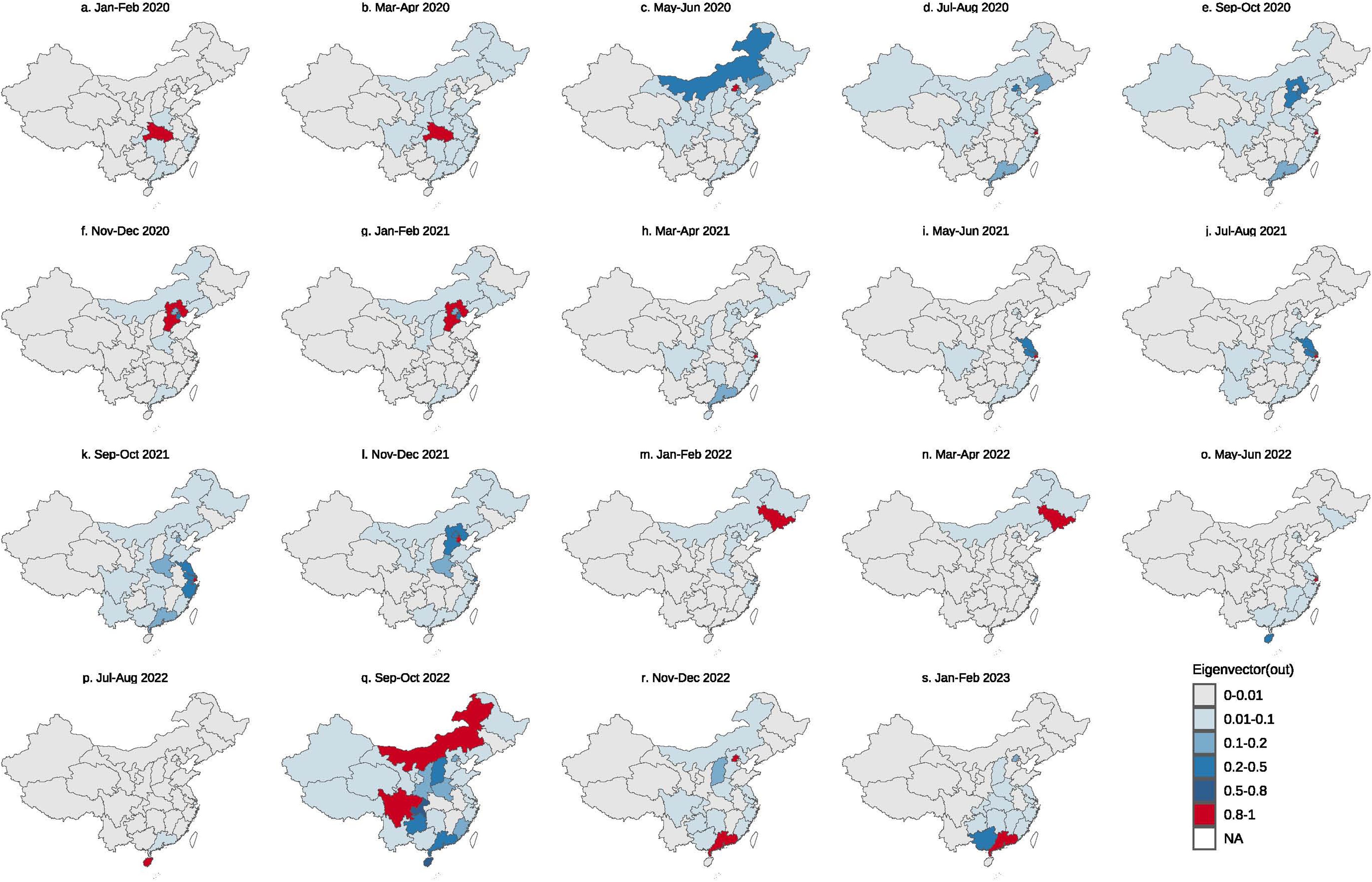
The spatiotemporal distribution of eigenvector-based key source locations in mainland China from January 2020 to February 2023, presented at a bimonthly resolution. Color gradients ranging from light to dark denote increasing importance of each location as a source, with corresponding values of the importance index indicated in the legend.

**Extended Data Fig. 4.**
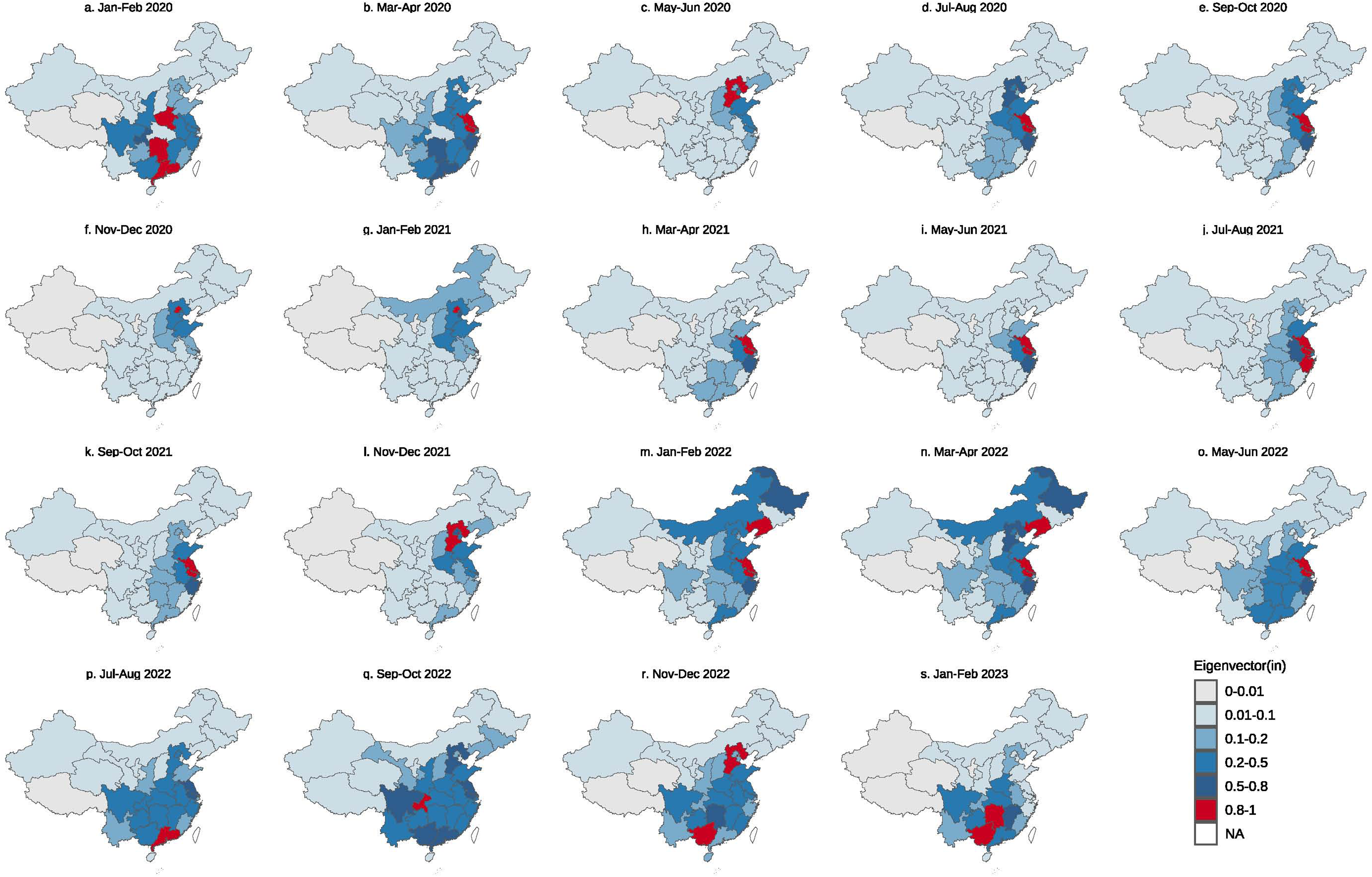
The spatiotemporal distribution of eigenvector-based vital sink locations in mainland China from January 2020 to February 2023 on a bimonthly scale. The color from light to dark indicates the importance of location from weak to strong, and the importance index in specific regions is shown in the legend.

**Extended Data Fig. 5.**
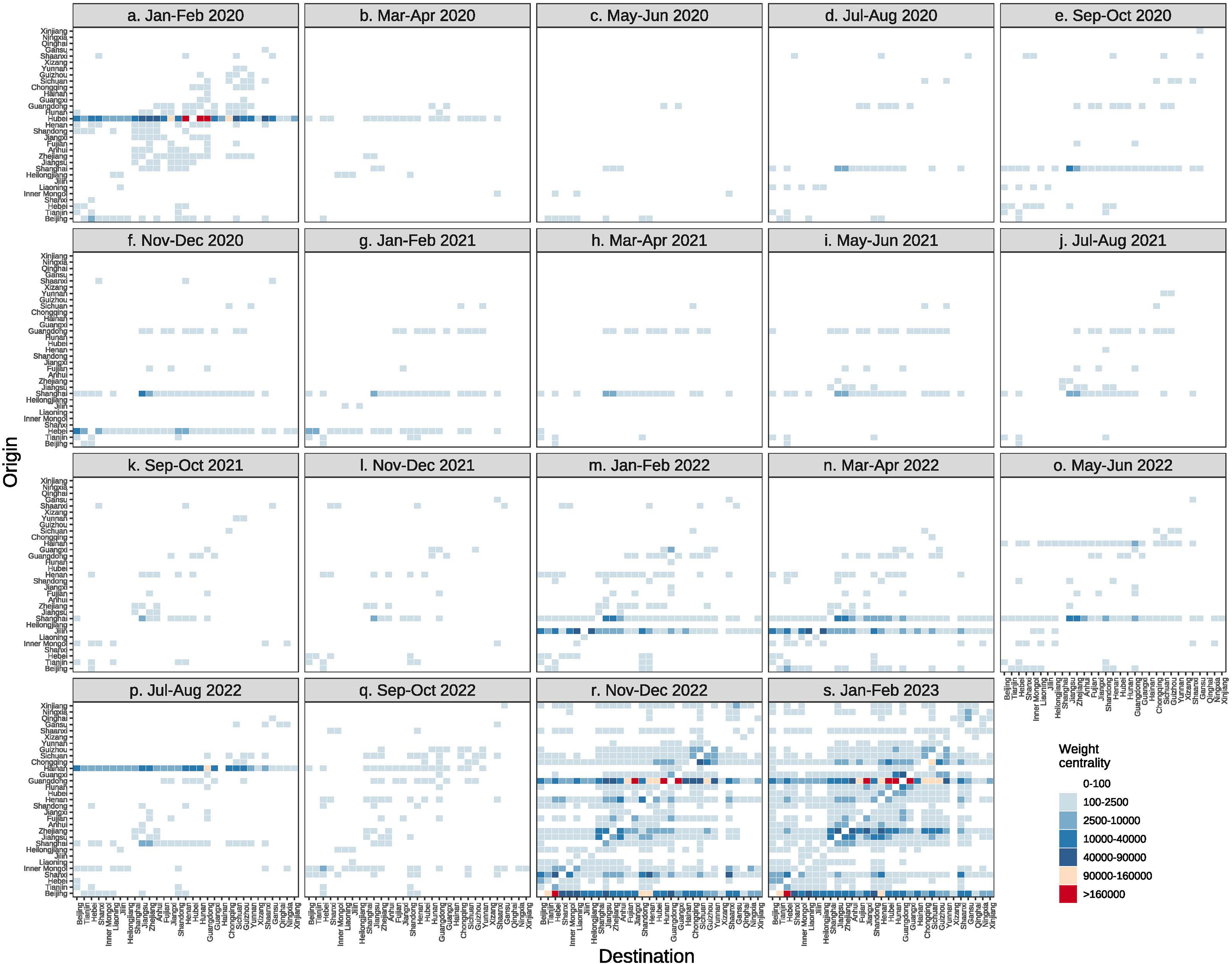
Key link properties of SARS-CoV-2 transmission networks in mainland China, quantified by link weight centrality at bimonthly intervals from January–February 2020 to January–February 2023. Higher centrality values denote greater significance within the network.

**Extended Data Fig. 6.**
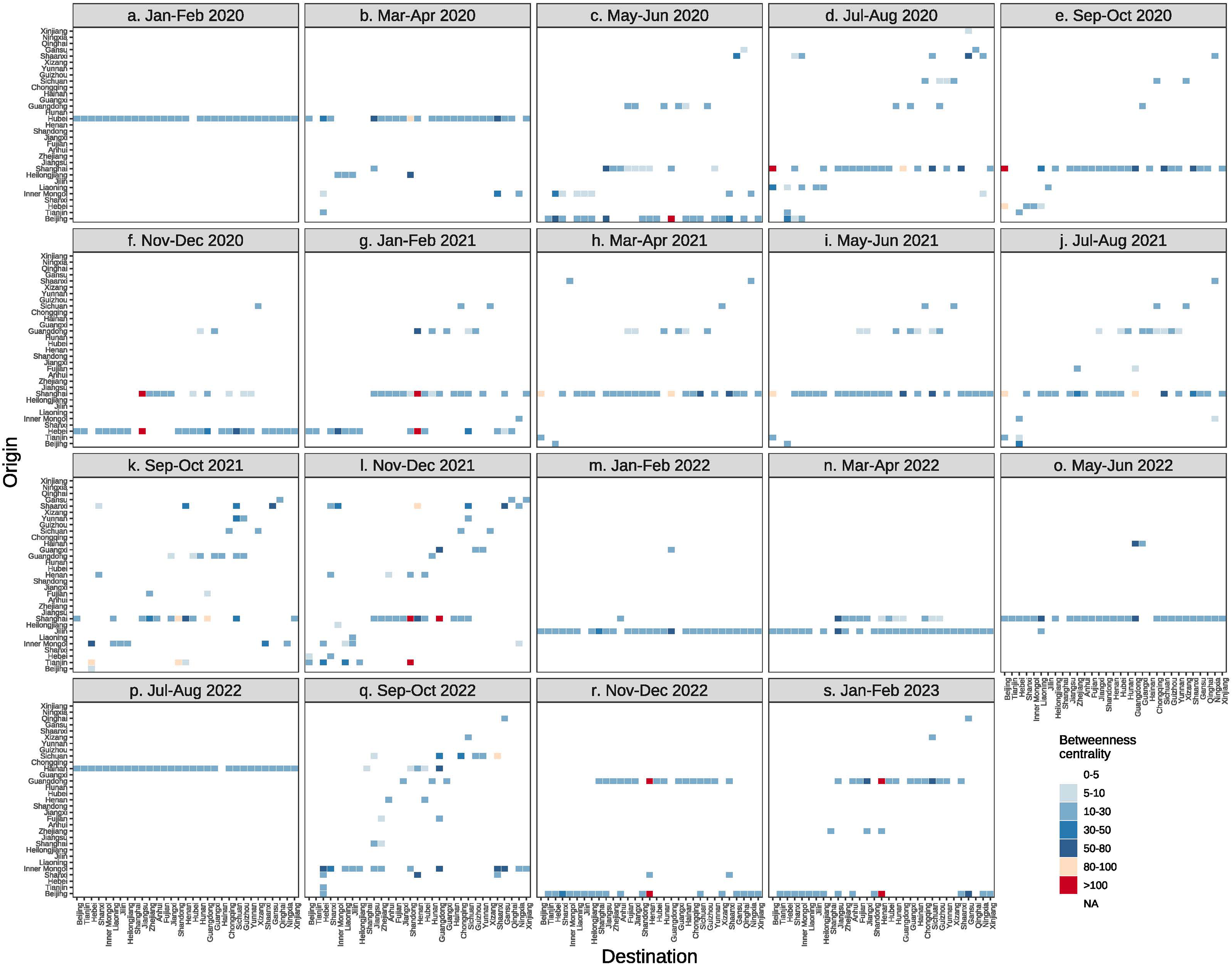
Key link properties of SARS-CoV-2 transmission networks in mainland China, quantified by betweenness centrality at bimonthly intervals from January–February 2020 to January–February 2023. Higher centrality values denote greater significance within the network.

**Extended Data Fig. 7.**
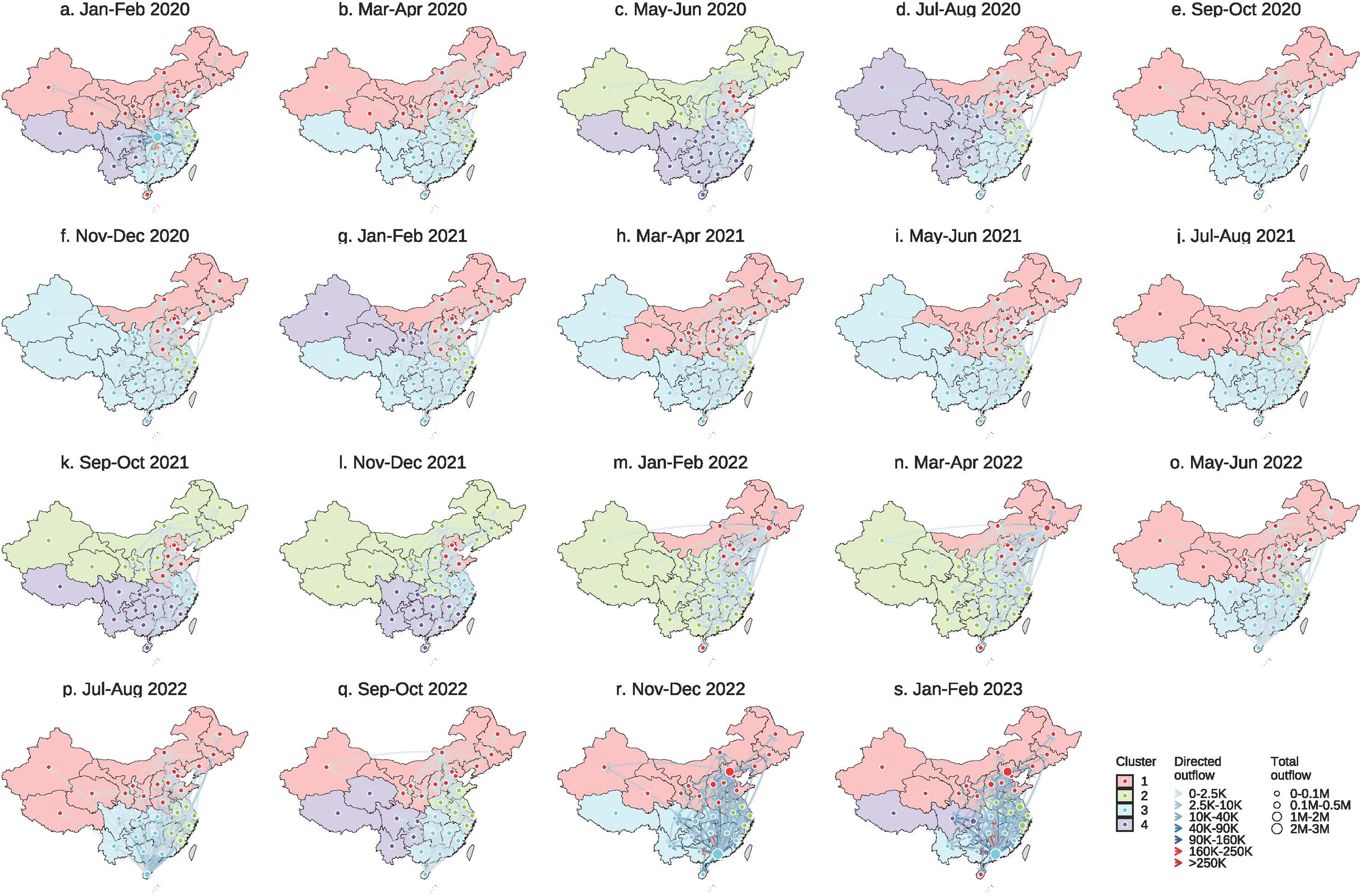
The geographic cluster distribution of the transmission network at a bimonthly scale from January 2020 to February 2023. The same colour in a subfigure denotes the same cluster of provinces. The node size represents the outflows of unreported infectious cases and the exposed population in that province. For each period, we showed the top 100 directed risk mobility.

**Extended Data Fig. 8.**
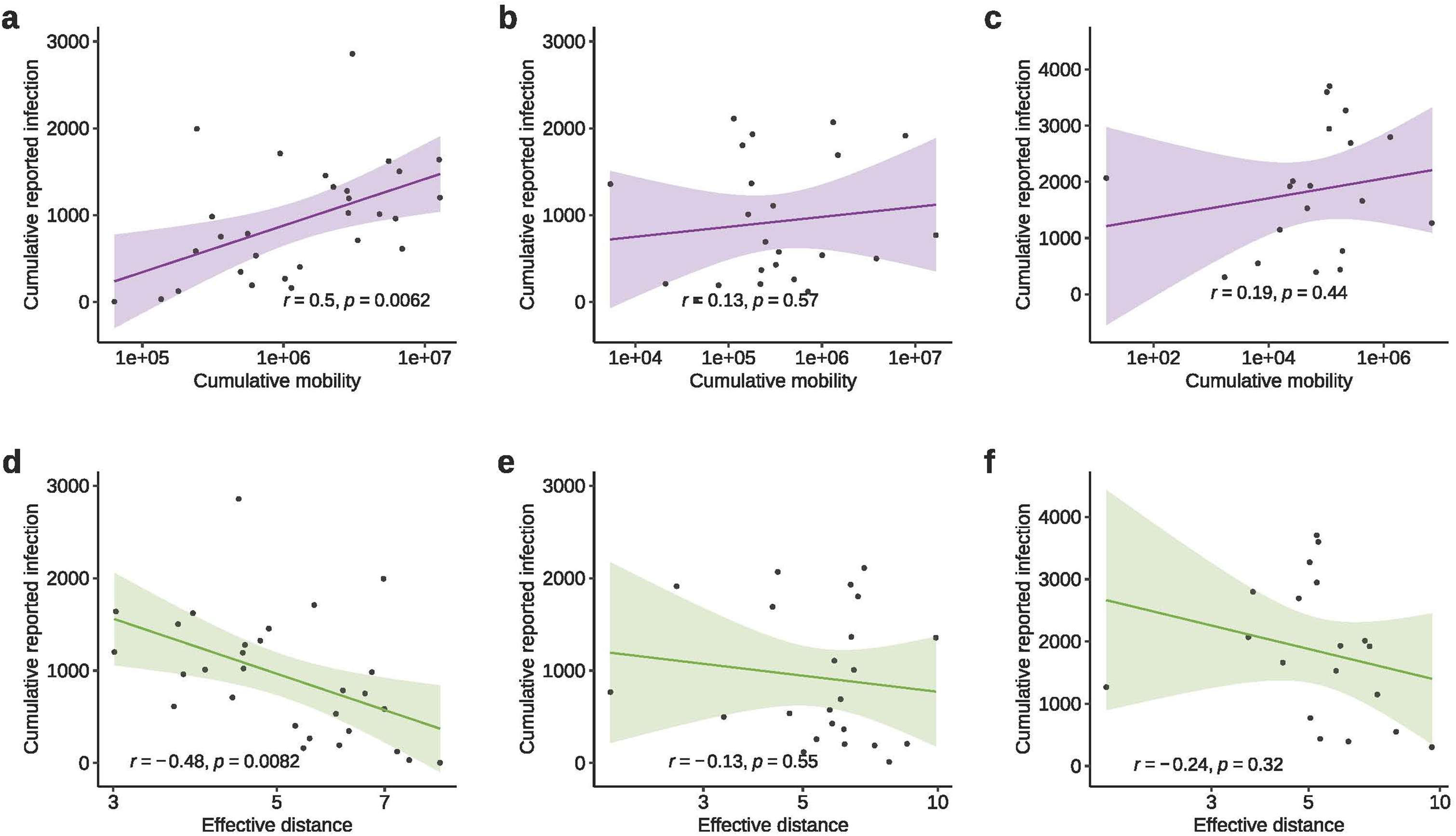
The associations between human mobility and SARS-CoV-2 transmission dynamics across three pandemic periods in mainland China from January 2020 to February 2023. (a, d) Relationships during the ancestral strain period: (a) reported cases versus inter-provincial mobility, (d) reported cases versus effective distance (b, e) and (c, f) present analogous analyses for the Alpha/Delta and Omicron variant periods, respectively.

**Extended Data Fig. 9.**
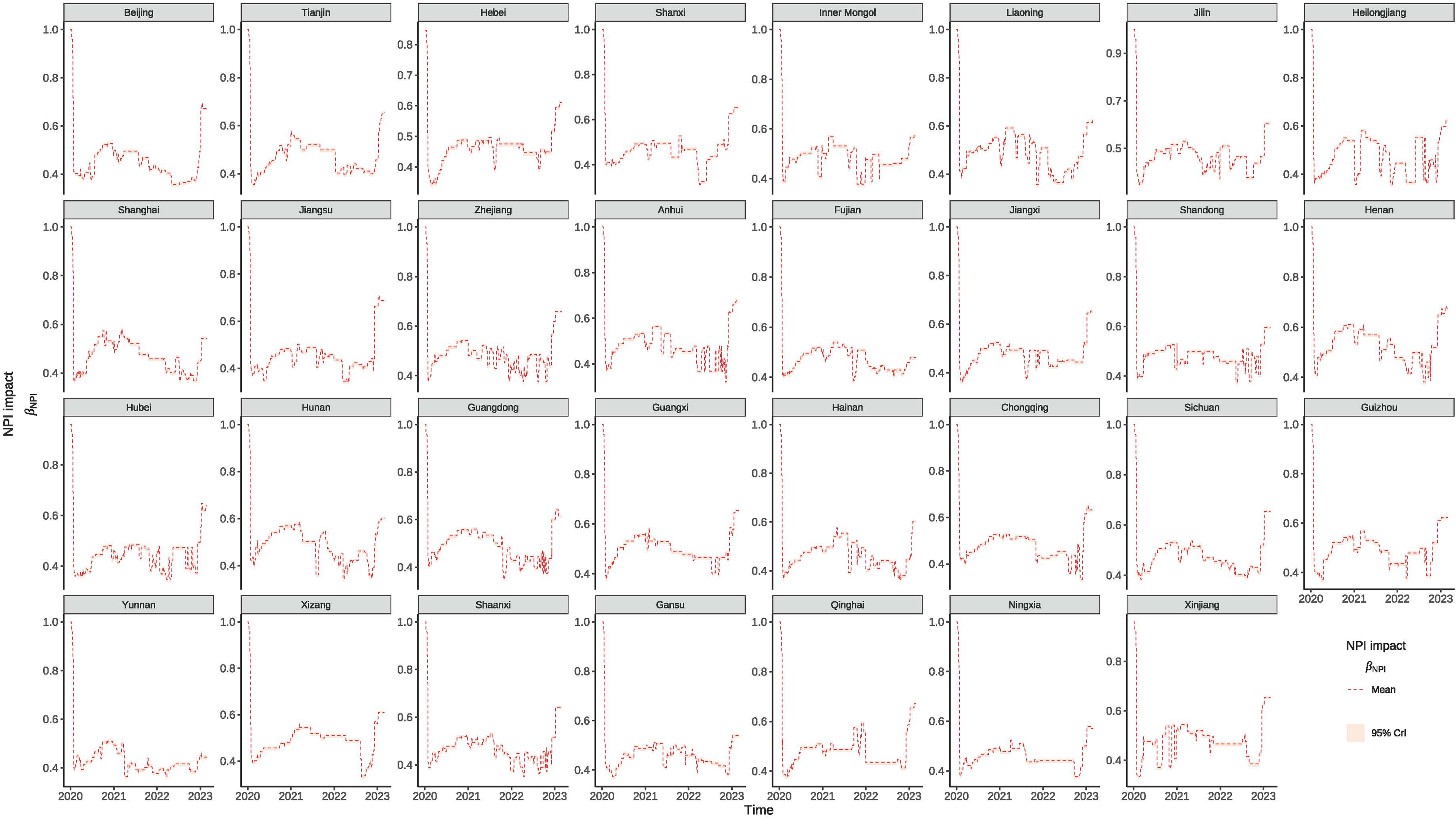
Temporal trends in the regional impacts of non-pharmaceutical interventions (NPIs) on COVID-19 transmission rate across mainland China from January 2020 to February 2023. Each section of the figure represents a different province.

**Extended Data Fig. 10.**
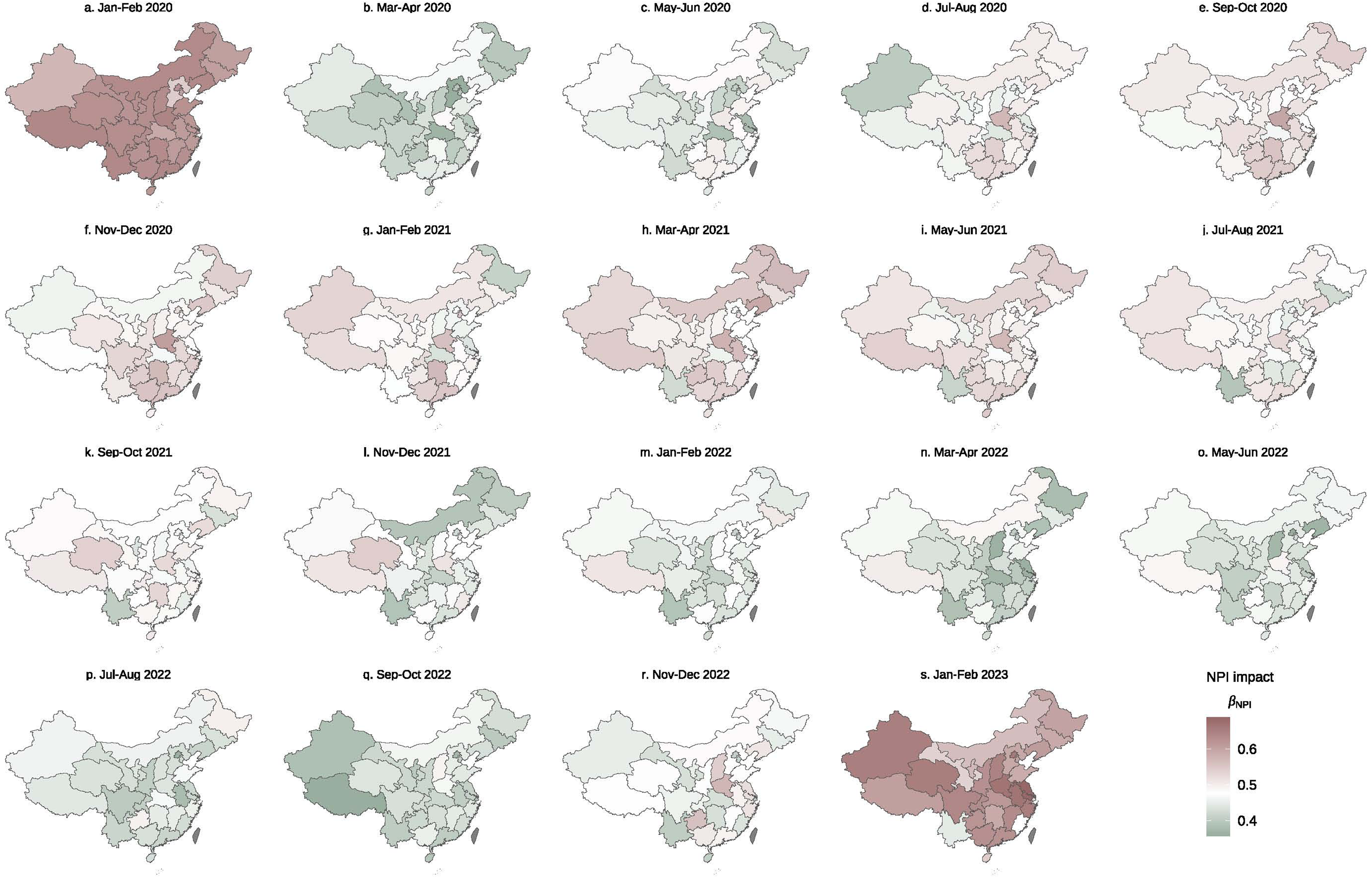
The spatiotemporal distribution of the impact of NPI on COVID-19 transmission rate in mainland China from January 2020 to February 2023 on a bimonthly scale. Color intensity, progressing from light to dark, corresponds to increasing impact levels, with quantitative values for specific regions detailed in the accompanying legend.

