## SUPPLEMENTARY MATERIALS for "Collective interactions, human mobility and viral evolution shaped the SARS-CoV-2 transmission in Mainland China"

of COVID-19 to endemicity very for Health, Hong Kong Science and Technology Park, Hong Kong Special Administrative Region, China

[3] School of Science, Harbin Institute of Technology, Shenzhen, China

[4] Interuniversity Institute for Biostatistics and Statistical Bioinformatics (I-BioStat), Data Science Institute, Hasselt University, Hasselt, Belgium

[5] Musketeers Foundation Institute of Data Science, The University of Hong Kong, Hong Kong, China

[6] Department of Pharmacology and Pharmacy, Li Ka Shing Faculty of Medicine, The University of Hong Kong, Hong Kong, China

### 1. Data sources and assimilation for the model

#### 1.1 Reported case Data for SARS-CoV-2

The epidemiological data utilized in this study comprise COVID-19 infection case counts reported from 31 provinces across 31 provinces mainland China. These data are organized as 31 distinct time series, each charting the temporal trajectory of infection cases within an individual province (Fig. S1). Furthermore, bimonthly infection maps are provided in Fig. S2, offering a comprehensive spatiotemporal perspective on transmission dynamics throughout the entirety of mainland China [10-12].

#### 1.2 SARS-CoV-2 genomic diversity data

Data on the SARS-CoV-2 variants of concern circulating across mainland China during the study period were obtained from publicly available genomic submissions to GISAID (https://gisaid.org/, GISAID: EPI_SET_250805xc), as illustrated in Extended Data Fig. 1. All genome sequences and associated metadata in this dataset are published in GISAID's EpiCoV database[1]. To view the contributors of each individual sequence with details such as accession number, Virus name, Collection date, Originating Lab, Submitting Lab, and the list of Authors, visit https://doi.org/10.55876/gis8.250805xc. These datasets enabled us to characterize the temporal dynamics of the emergence and predominance of variants of concern over the course of the investigation.

#### 1. 3 Sequence acquisition and quality control

The reference nucleotide sequence of the SARS-CoV-2 Spike glycoprotein gene (GenBank accession: NC_045512.2; genomic coordinates 21,563–25,384, comprising 3,822 nucleotides) was obtained from the NCBI database (https://www.ncbi.nlm.nih.gov/nuccore/NC_045512.2/). Comprehensive sets of viral genomic sequences encompassing the Spike gene were procured from the GISAID EpiCoV database (https://www.gisaid.org/). Robust quality control measures were employed, restricting inclusion to sequences sampled within China and collected between 1 January 2020 and 28 February 2023. Eligible sequences were further required to exhibit a length within ±2% of the reference (3,746–3,898 nucleotides), to contain fewer than 5% ambiguous bases (N) with no consecutive stretch of 10 or more, to possess more than 95% canonical nucleotide characters (A, C, G, T), and to lack internal stop codons in the Spike gene open reading frame [2,3]. Only sequences meeting all specified criteria were retained for subsequent analyses and consolidated into a FASTA file.

#### 1.4 Sequence alignment and genetic diversity analysis

Multiple sequence alignment of the high-confidence Spike gene sequences was performed via MAFFT (v7.526) using default parameters [4]. The degree of genetic diversity within the viral cohort was quantified by estimating nucleotide diversity (π), defined as the mean number of pairwise nucleotide differences per site. This parameter was calculated with the nuc.div() function from the pegas package (v1.1) in the R statistical environment (v4.0.5) [5]. To elucidate temporal trends, nucleotide diversity (π) was calculated on a monthly basis by aggregating sequences by collection month, thereby enabling a comprehensive assessment of the temporal evolution of Spike gene diversity in the Chinese SARS-CoV-2 population across the duration of the study.

##### 1.5 Antigenic drift analysis

To obtain the antigenic drift data, the receptor-binding domain (RBD, 318–541) was extracted from each aligned sequence [6,7]. Antigenic drift was quantified as the mean p-distance between RBD sequences from the current month and those from the previous three months[8,9], computed using a block-matrix approach to estimate pairwise amino acid differences.

##### 1.6 NPI stringency Data

In this study, we incorporate a range of auxiliary datasets, including daily indices of Non-Pharmaceutical Interventions (NPIs), as depicted in Figs. S3–S4. These indices were obtained from the Oxford COVID-19 Government Response Tracker (OxCGRT) (https://github.com/OxCGRT/covid-policy-tracker) and quantify the stringency of public health and social measures enforced to mitigate the transmission of COVID-19 [13].

##### 1.7 Outcomes of time-varying characteristics and their distributions for key sources and sinks

We characterize the spatiotemporal distribution of key sources and sinks[14,15], by analyzing multiple centrality measures—including degree centrality, eigenvector centrality, hub scores, and PageRank centrality—within the reconstructed transmission network. While Fig. 3 in the main text highlights prominent locations based on eigenvector centrality, here we extend this analysis to other centrality metrics, as presented in Figs. S5–S7. Furthermore, we elucidate the dynamic properties of critical sources and sinks by providing bimonthly, temporally resolved maps depicting their evolution across various phases of the pandemic (Figs. S8–S10 and S11–S13, respectively).

##### 1.8 Time-varying patterns in essential transmission pathways

To elucidate the temporal dynamics of pivotal transmission routes, we assess link weight centrality and betweenness centrality, as depicted in Extended Data Fig. 5 and Extended Data Fig. 6, respectively. These network metrics allow us to identify the principal conduits underpinning viral dissemination. In the early stage of the pandemic, dominant transmission pathways originated from Hubei, extending chiefly towards Henan and Guangdong. During the intermediate phase, the primary axis of transmission shifted to the Shanghai–Jiangsu corridor. In the final phase, the network structure became increasingly diffuse, with multiple links emerging as significant pathways for viral spread.

##### 1.9 Temporal evolution of geographic clusters

In parallel with Fig. 5 of the main text, which illustrates geographic cluster during three distinct phases characterized by variants of concern, we present here the dynamics of clustering patterns at a bimonthly resolution (Extended Data Fig. 7). This higher temporal granularity affords a more nuanced view of the evolving geographic cluster structures throughout the course of the pandemic.

#### 1.10 Time-varying transmissibility

As the measure of time-varying transmissibility, we opted the effective reproductive number, commonly denoted as $R_e$, quantifies the mean number of secondary infections generated by a single infected individual at time $t$. This parameter is pivotal for elucidating the transmission dynamics of COVID-19 and for evaluating the influence of various determinants on viral spread. The daily trajectories of $R_e$, as illustrated in Fig. S14, offer insights into temporal variations in transmissibility in relation to implemented interventions, which is comparable with previous references [16,17].

To further capture the spatiotemporal variation of $R_e$, we provide bimonthly maps displaying the effective reproductive number across provinces (Fig. S15). These visualizations underscore pronounced regional disparities and evolving temporal patterns in transmission potential, thereby affording a comprehensive overview of the spatial and temporal variability in COVID-19 transmissibility across mainland China.

#### 1.11 Ascertainment rate

The ascertainment rate represents the proportion of actual COVID-19 cases that are identified and formally reported[11]. This parameter is vital for accurately gauging the true scale of viral spread and evaluating the effectiveness of diagnostic and surveillance systems. The temporal trajectories of the ascertainment rate, as depicted in Fig. S16, elucidate how case detection fluctuates over time in response to NPIs and other influencing factors.

To further characterize the spatiotemporal variation of the ascertainment rate, we present bimonthly maps illustrating its variation across provinces (Fig. S17). These visualizations reveal marked regional disparities and temporal shifts in case detection and reporting, thus providing a comprehensive perspective on the evolution of the ascertainment rate across both spatial and temporal dimensions.

#### 2. Supplementary tables

##### Table S1. Descriptions of model parameters and their estimations for reliable choice of prior distributions.

| *Parameters* | *Description* | *Value/Prior distribution* | *References* | *Estimates* |
|---|---|---|---|---|
| $1/\sigma$ | The mean latent period of the infected individuals (days) | 5 days | [11,18,19] | - |
| $1/\gamma$ | The infectious period of infected individuals (days) | 10 days | [11,18,19] | - |
| $1/q$ | The duration of influenza immunity from infection (days) | 610 days | [20] | - |
| $p_i$ | The proportion of infections influenced by social reinforcement under the higher-order collective interactions for province *i* | HalfNormal (0,1) | Model estimates | See Extended Data Fig. 2 |
| $\phi_i$ | The impact of social reinforcements on the susceptibility for province *i* | HalfNormal (1,5) | Model estimates | See Extended Data Fig. 3 |
| $v_0(t)$ | The baseline impact of social reinforcements on the transmissibility during period *t* | HalfNormal (0,1) | Model estimates | See Fig 2(c) |
| $\alpha_1$ | A coefficient determines the strength of the impact of diversity on transmissibility. | HalfNormal (0,1) | Model estimates | 0.64 (0.02,2.17) |
| $\alpha_2$ | A coefficient determines the strength of the impact of antigenic drift on susceptibility. | HalfNormal (0,1) | Model estimates | 0.07 (0.05, 0.09) |
| $\pi_1$ | A coefficient determines the strength of the impact of NPI on transmission rate. | HalfNormal (0,5) | Model estimates | 1.33 (1.31, 1.37,) |
| $\pi_2$ | A coefficient determines the strength of the impact of NPI on ascertainment rate. | HalfNormal (1,10) | Model estimates | 9.58 (9.15,10.00) |

### 3. Supplementary figures

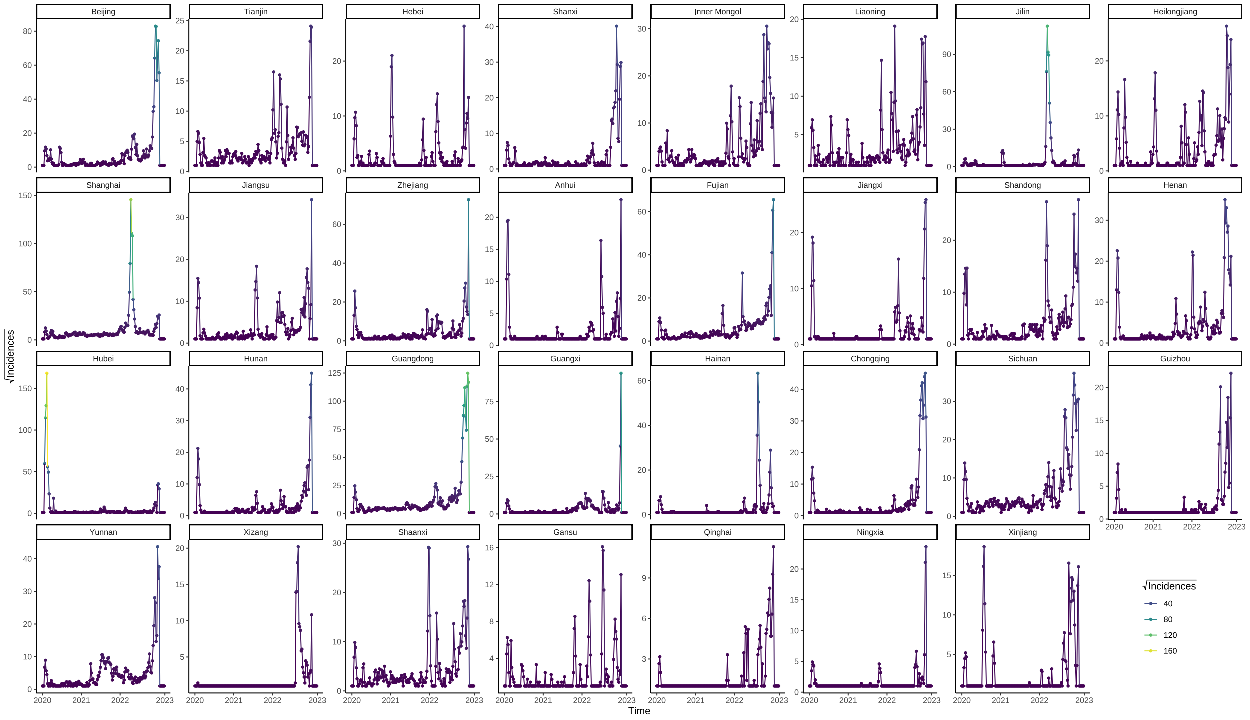


**Figure S1**. Temporal trends in regional COVID-19 incidence across mainland China from January 2020 to February 2023, presented on a square root–transformed scale. Each panel corresponds to a distinct province, with numerals denoting the square root of reported case counts.

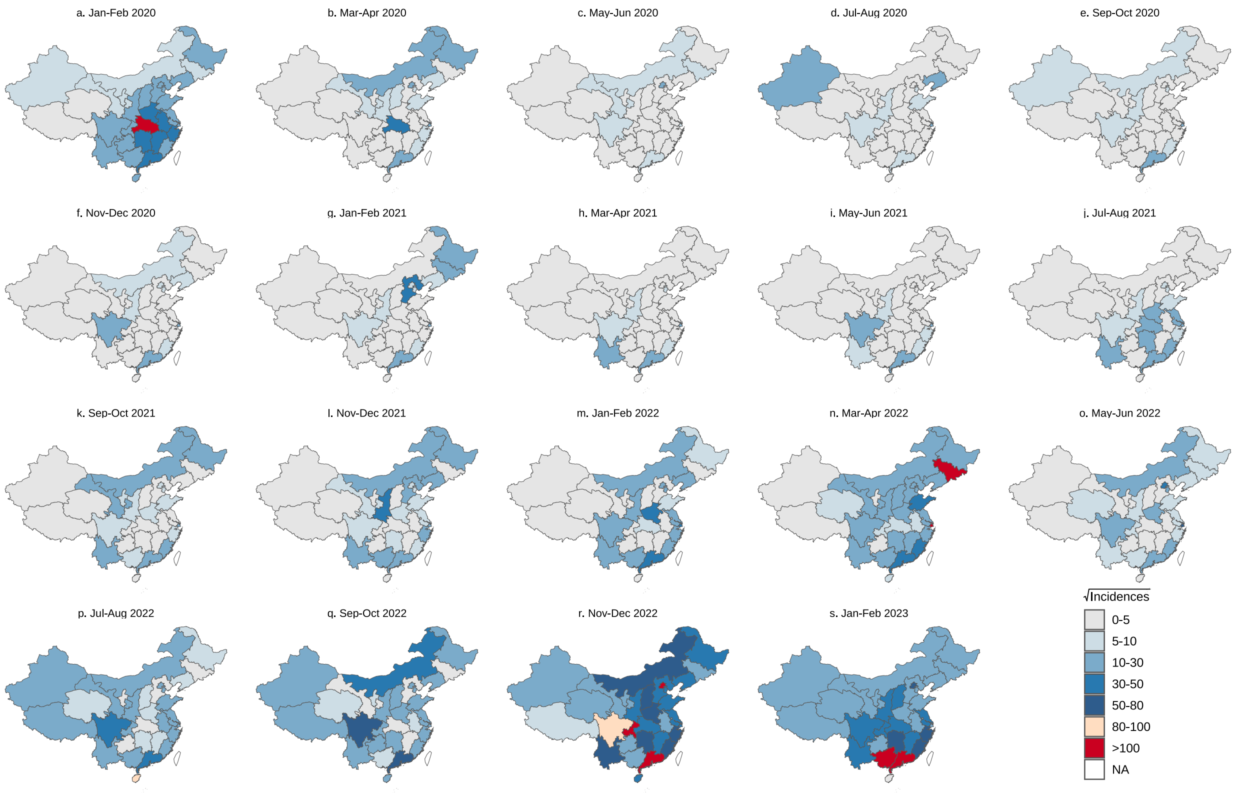


**Figure S2**. Spatiotemporal distribution of reported COVID-19 cases in mainland China from January 2020 to February 2023. Each section of the figure represents a different province, with numerals denoting the square root of reported case counts indicated in the legend.

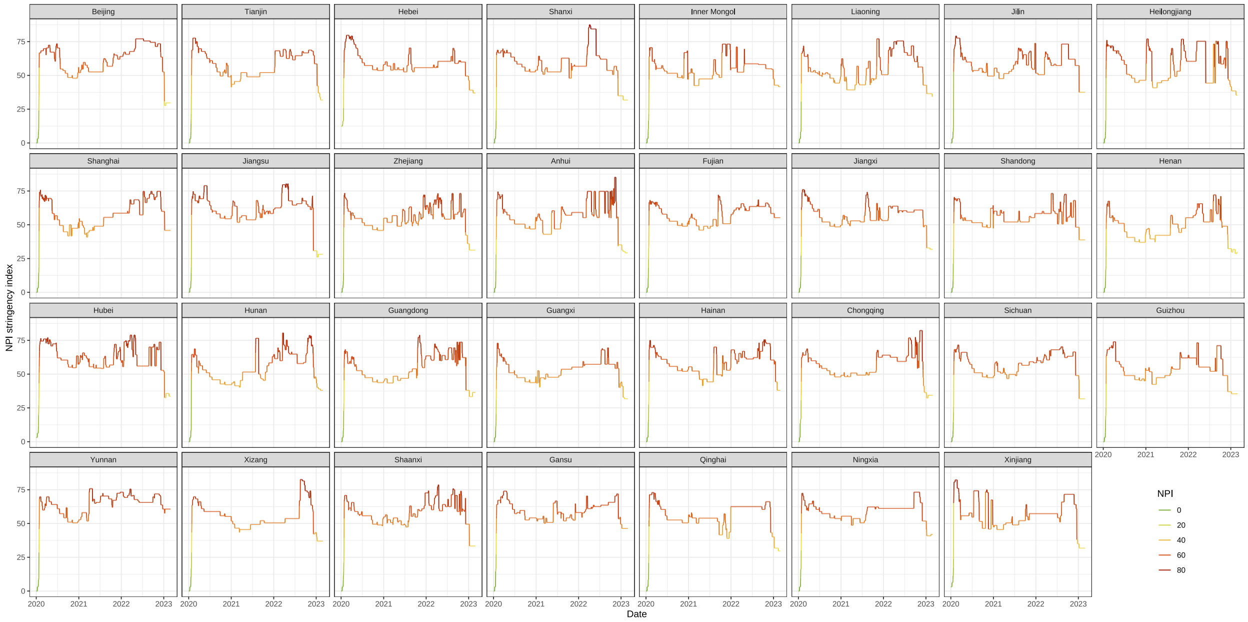


**Figure S3**. Regional trajectories of the OxCGRT non-pharmaceutical intervention (NPI) index in mainland China from January 2020 to February 2023. Each panel corresponds to a specific province, with numerals indicating the intensity of NPI measures over time.

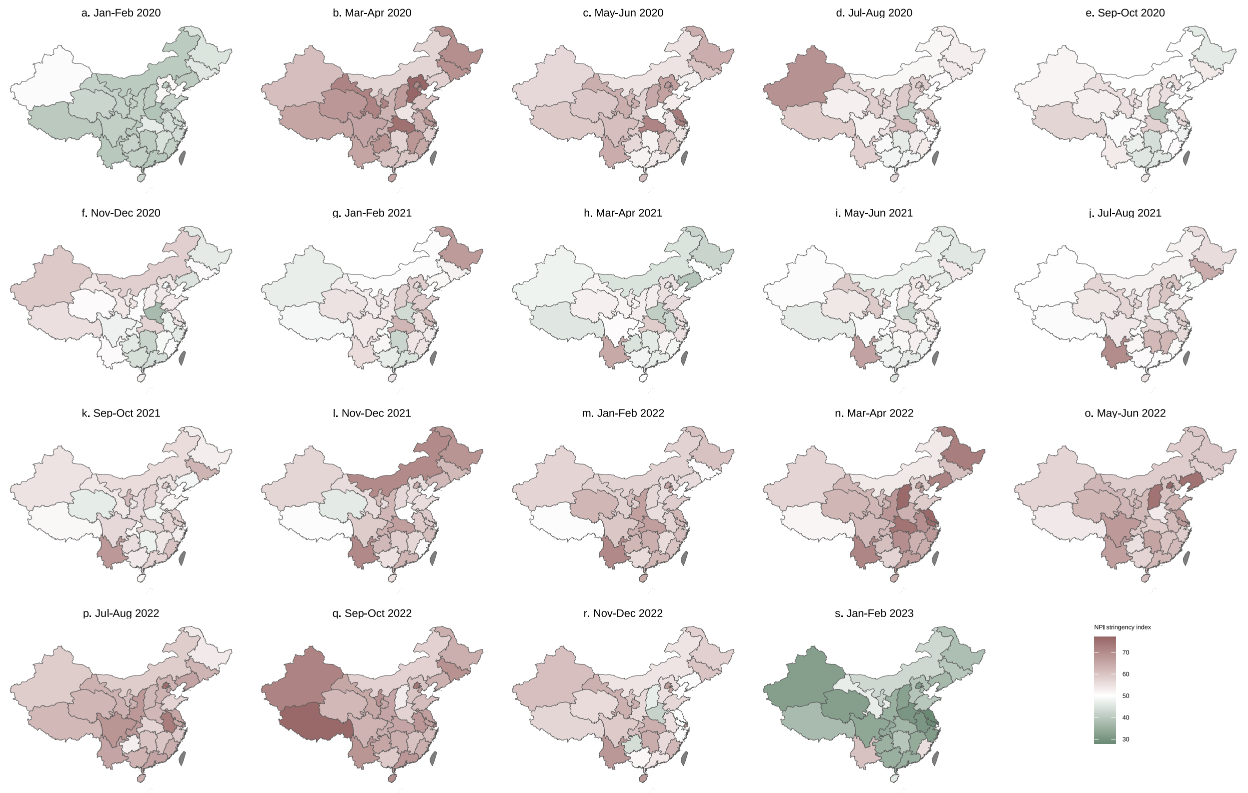


**Figure S4**. Spatiotemporal distribution of the OxCGRT NPI index across mainland China from January 2020 to February 2023. Color gradients, ranging from light to dark, represent increasing NPI stringency, with specific index values delineated in the accompanying legend.

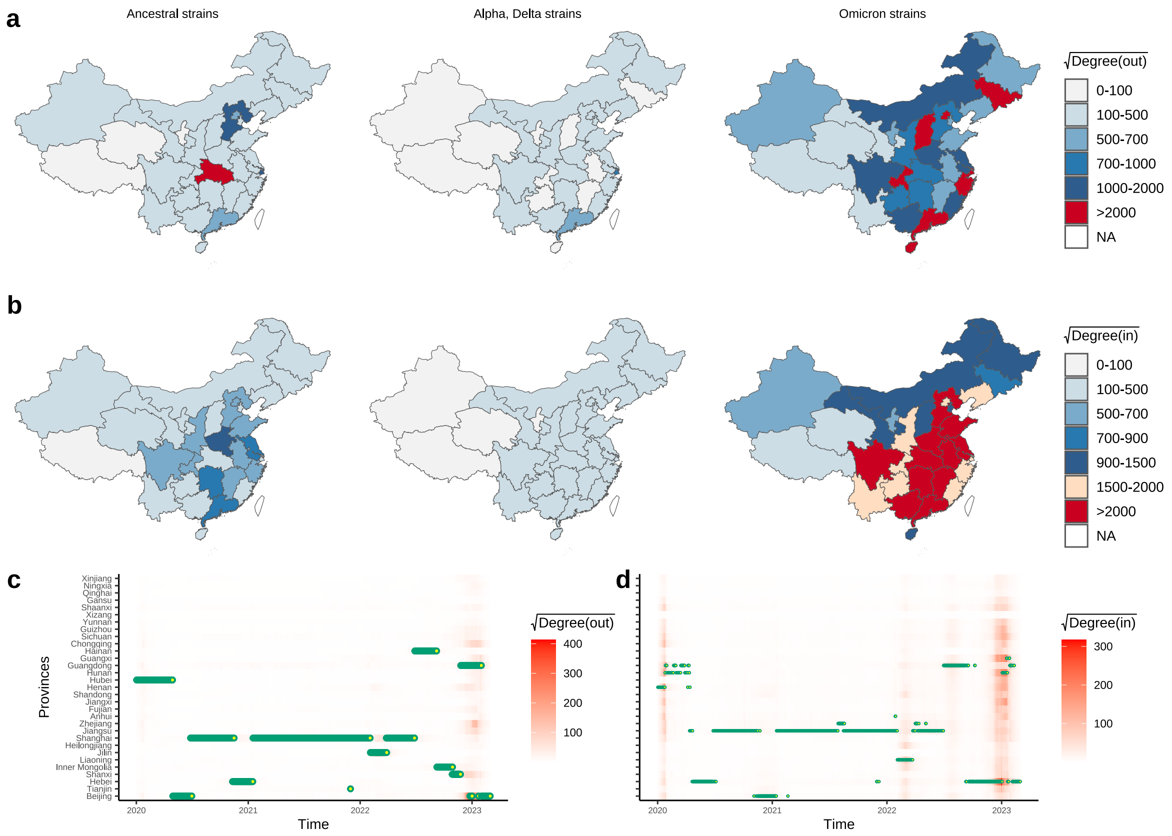


**Figure S5**. Identification of key transmission locations for SARS-CoV-2 in mainland China, from January 2020 to February 2023. (a) Spatial distribution of origin locations, as determined by degree centrality within the outgoing transmission network, highlighting provinces with the highest outflow of SARS-CoV-2 for the three major variants of concern. (b) Spatial distribution of key destination locations, based on eigenvector centrality for incoming transmission, indicating provinces with the highest inflow of SARS-CoV-2 associated with the three variants of concern. (c) Temporal evolution of dominant source (outflow) provinces identified as vital transmission origins. (d) Temporal evolution of major sink (inflow) provinces representing crucial destinations for viral spread. Greater intensity of red in the index corresponds to increasingly significant locations, while blue specifically denotes the most critical transmission hubs at each time point.

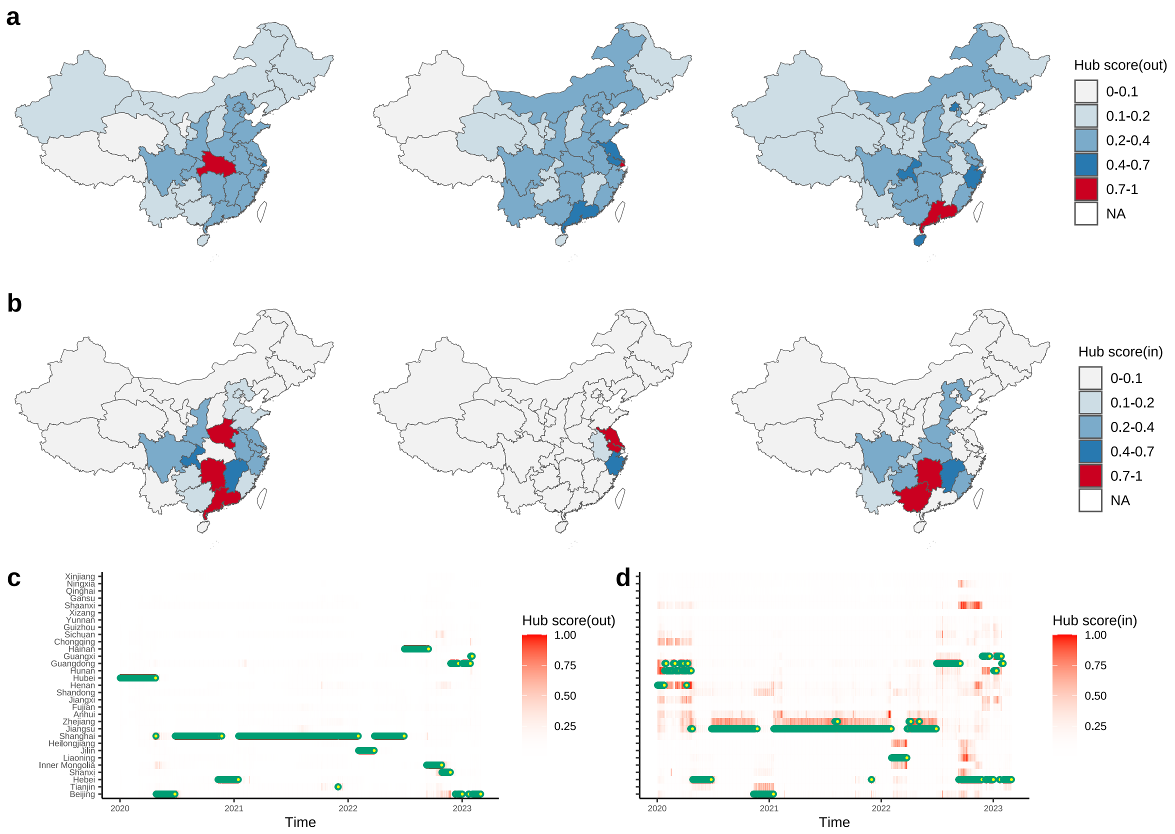


**Figure S6**. Identification of key transmission locations for SARS-CoV-2 in mainland China, from January 2020 to February 2023. (a) Spatial distribution of origin hubs, as determined by hub centrality within the outgoing transmission network, highlighting provinces with the highest outflow of SARS-CoV-2 for the three major variants of concern. (b) Spatial distribution of key destination locations, based on hub centrality for incoming transmission, indicating provinces with the highest inflow of SARS-CoV-2 associated with the three variants of concern. (c) Temporal evolution of dominant source (outflow) provinces identified as vital transmission origins. (d) Temporal evolution of major sink (inflow) provinces representing crucial destinations for viral spread. Greater intensity of red in the index corresponds to increasingly significant locations, while blue specifically denotes the most critical transmission hubs at each time point.

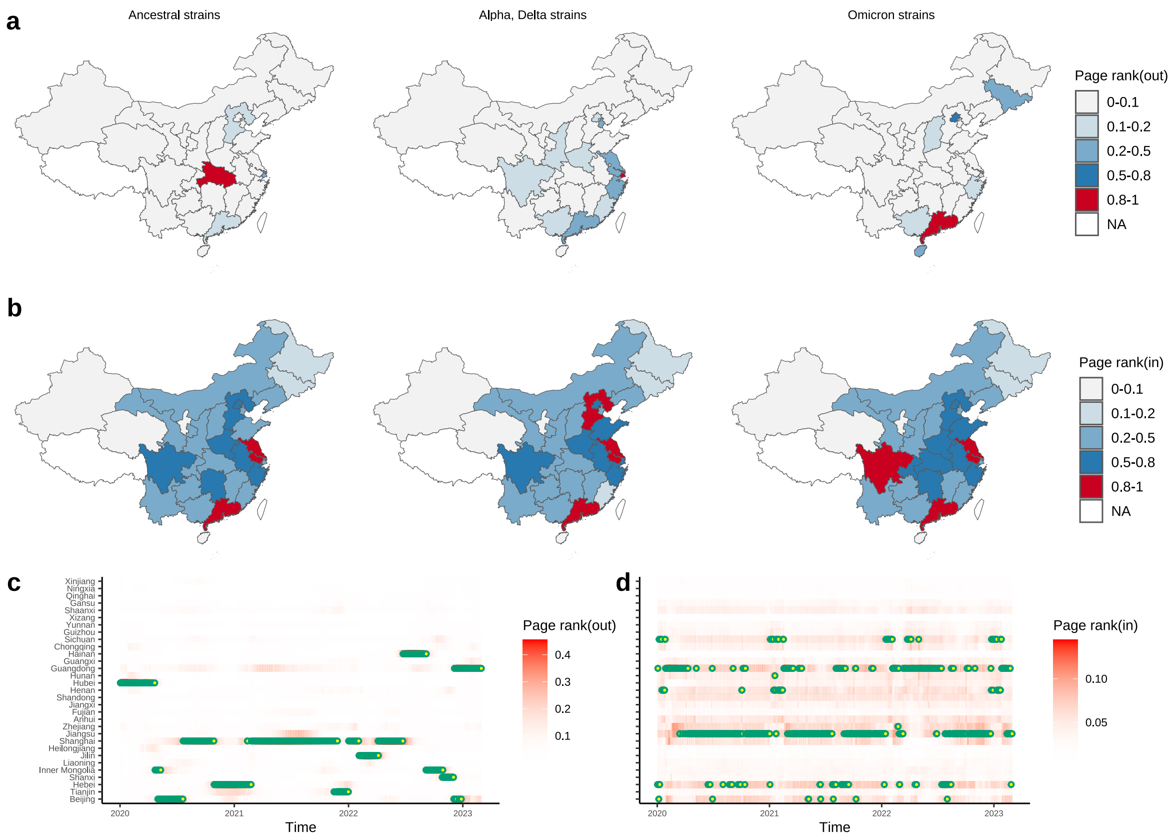


**Figure S7**. Identification of key transmission locations for SARS-CoV-2 in mainland China, from January 2020 to February 2023. (a) Spatial distribution of origin hubs, as determined by pagerank centrality within the outgoing transmission network, highlighting provinces with the highest outflow of SARS-CoV-2 for the three major variants of concern. (b) Spatial distribution of key destination locations, based on pagerank centrality for incoming transmission, indicating provinces with the highest inflow of SARS-CoV-2 associated with the three variants of concern. (c) Temporal evolution of dominant source (outflow) provinces identified as vital transmission origins. (d) Temporal evolution of major sink (inflow) provinces representing crucial destinations for viral spread. Greater intensity of red in the index corresponds to increasingly significant locations, while blue specifically denotes the most critical transmission hubs at each time point.

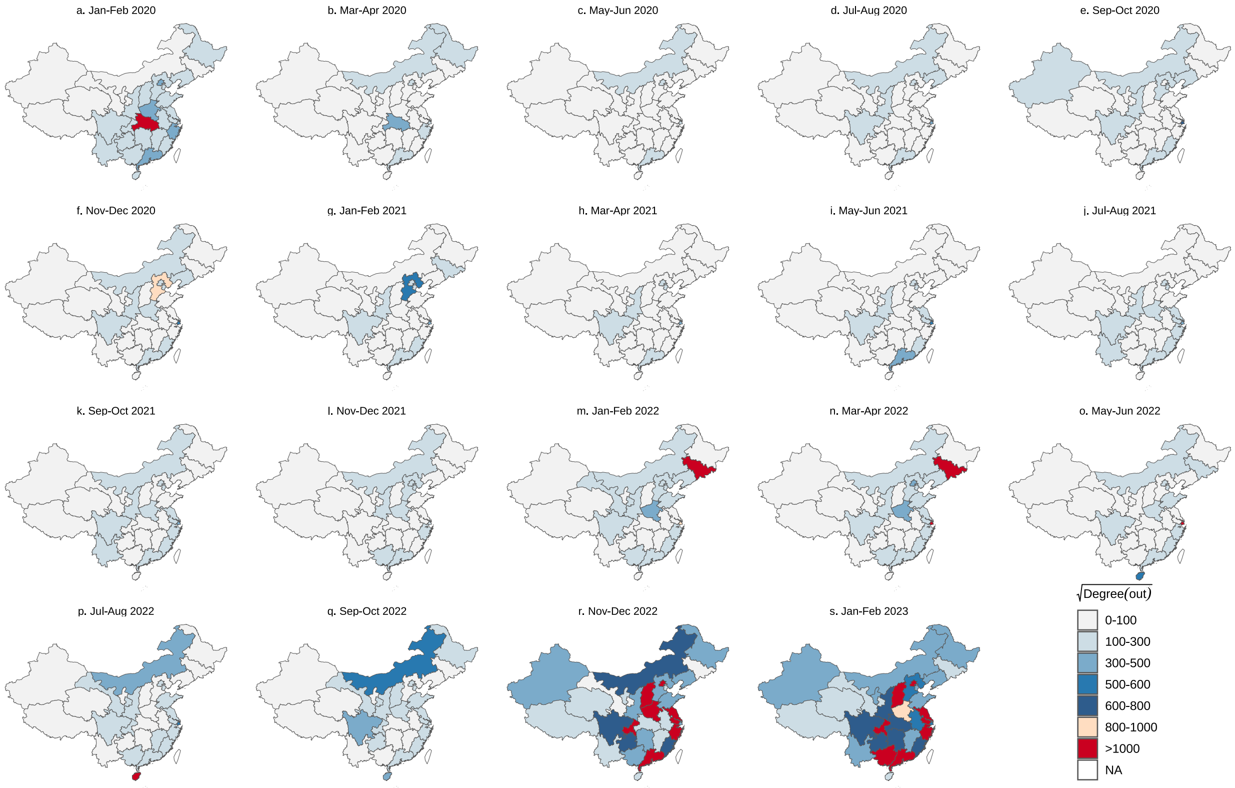


**Figure S8**. Spatiotemporal distribution of degree-based key source locations in mainland China from January 2020 to February 2023, presented at a bimonthly resolution. Color gradients ranging from light to dark denote increasing importance of each location as a source, with corresponding values of the importance index indicated in the legend.

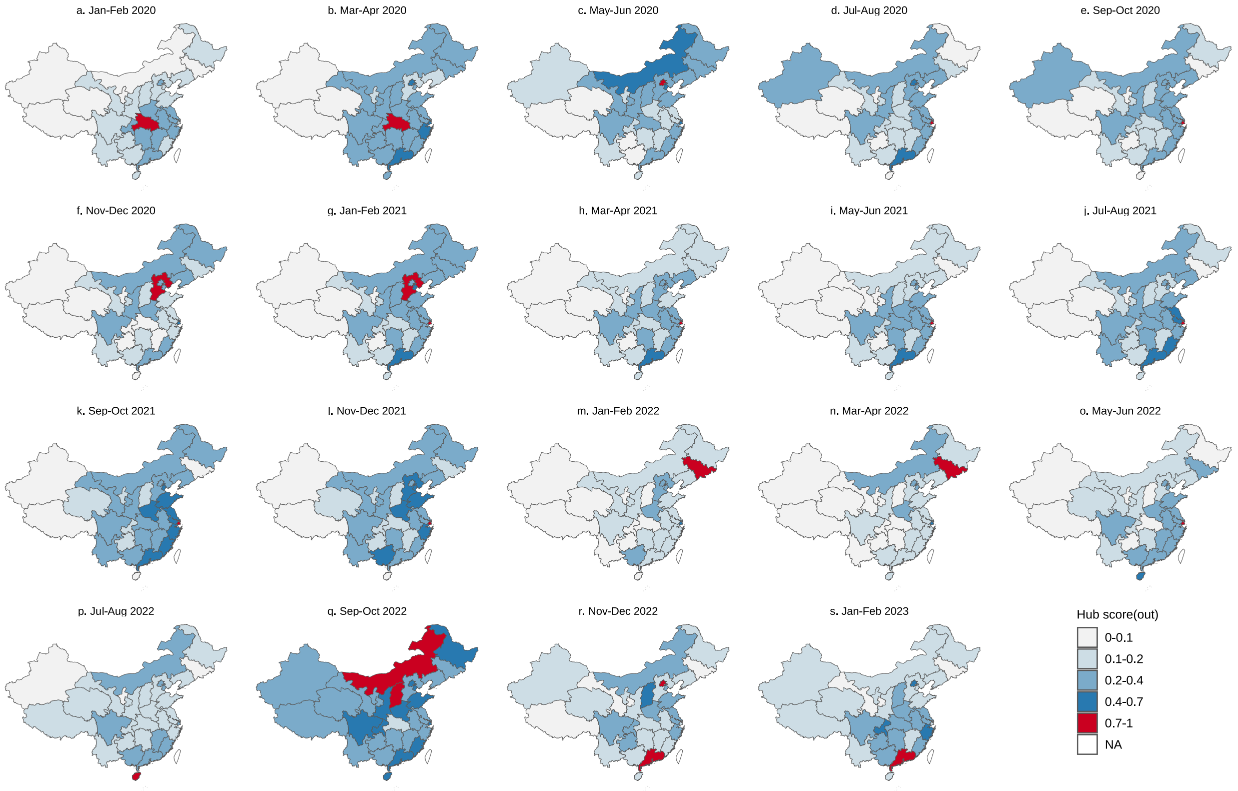


**Figure S9**. The spatiotemporal distribution of hub-score-based vital source locations in mainland China from January 2020 to February 2023 on a bimonthly scale. The color from light to dark indicates the importance of location from weak to strong, and the importance index in specific regions is shown in the legend.

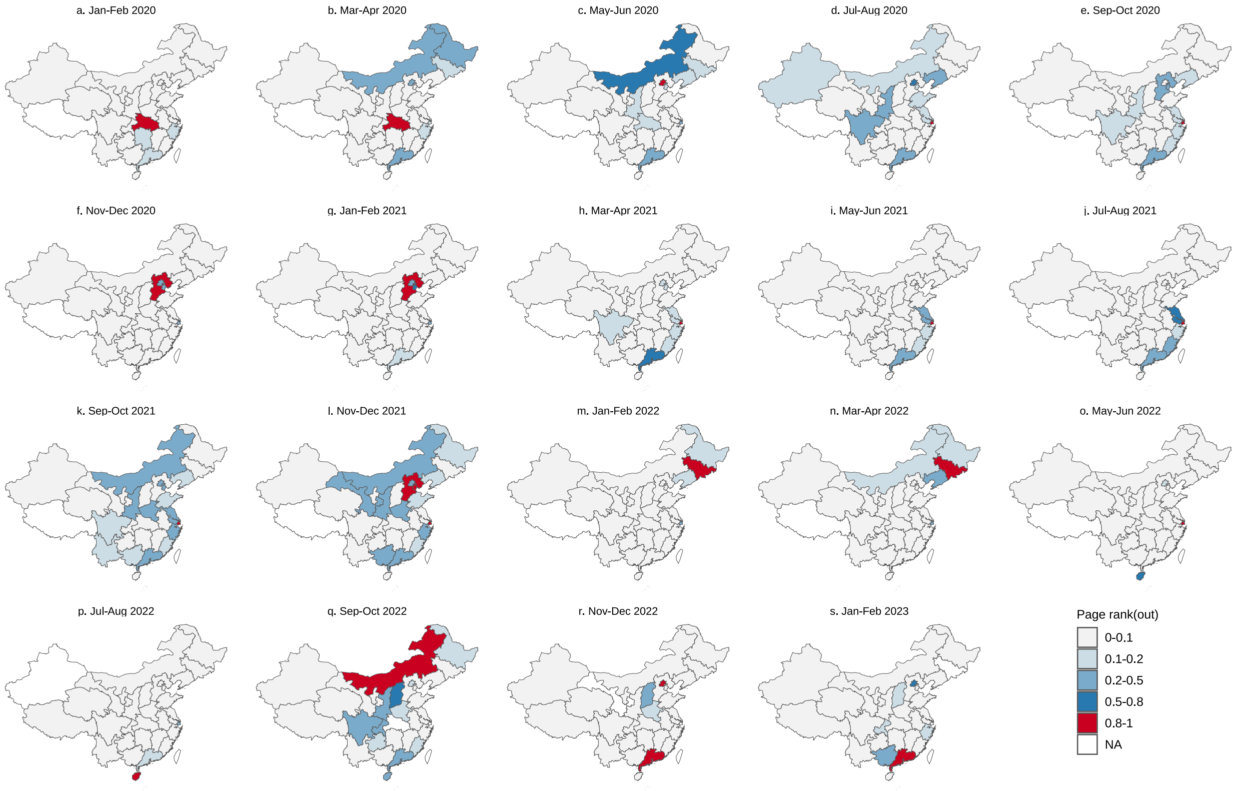


**Figure S10**. The spatiotemporal distribution of page-rank-based vital source locations in mainland China from January 2020 to February 2023 on a bimonthly scale. The color from light to dark indicates the importance of location from weak to strong, and the importance index in specific regions is shown in the legend.

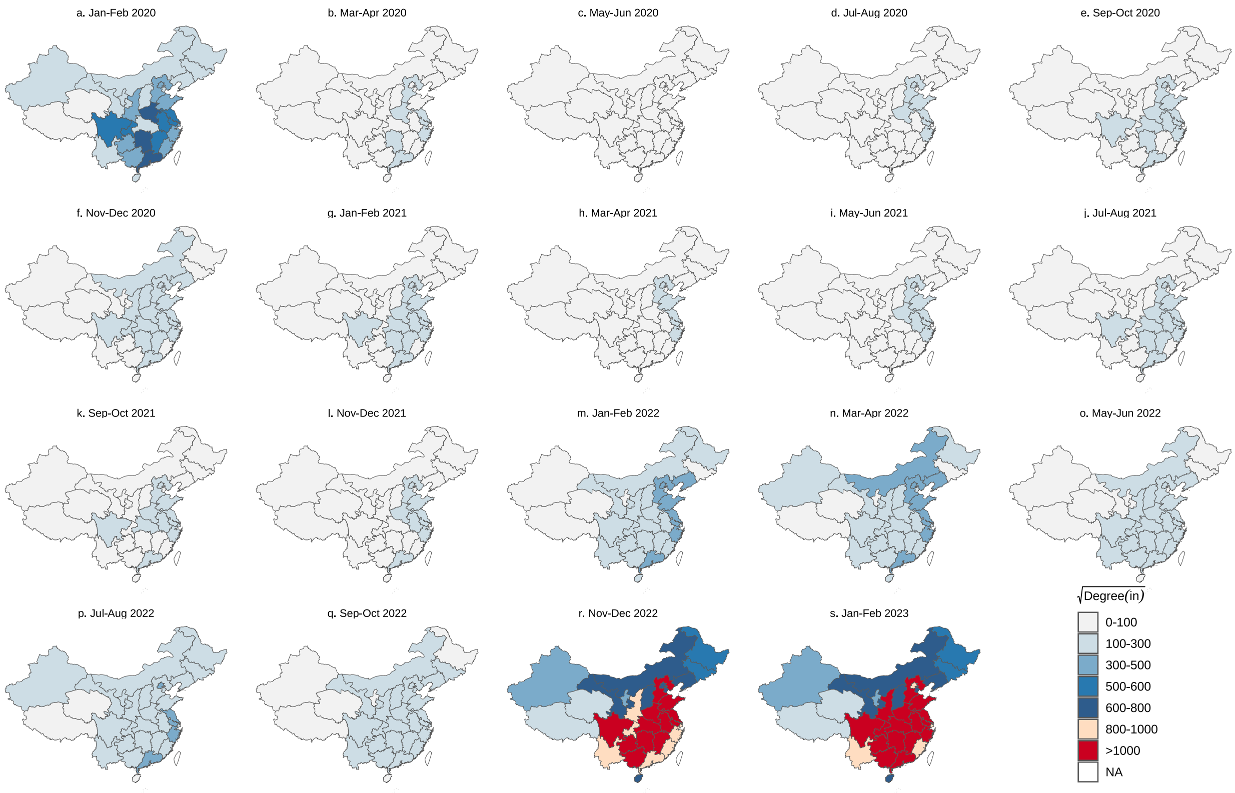


**Figure S11**. The spatiotemporal distribution of degree-based vital sink locations in mainland China from January 2020 to February 2023 on a bimonthly scale. The color from light to dark indicates the importance of location from weak to strong, and the importance index in specific regions is shown in the legend.

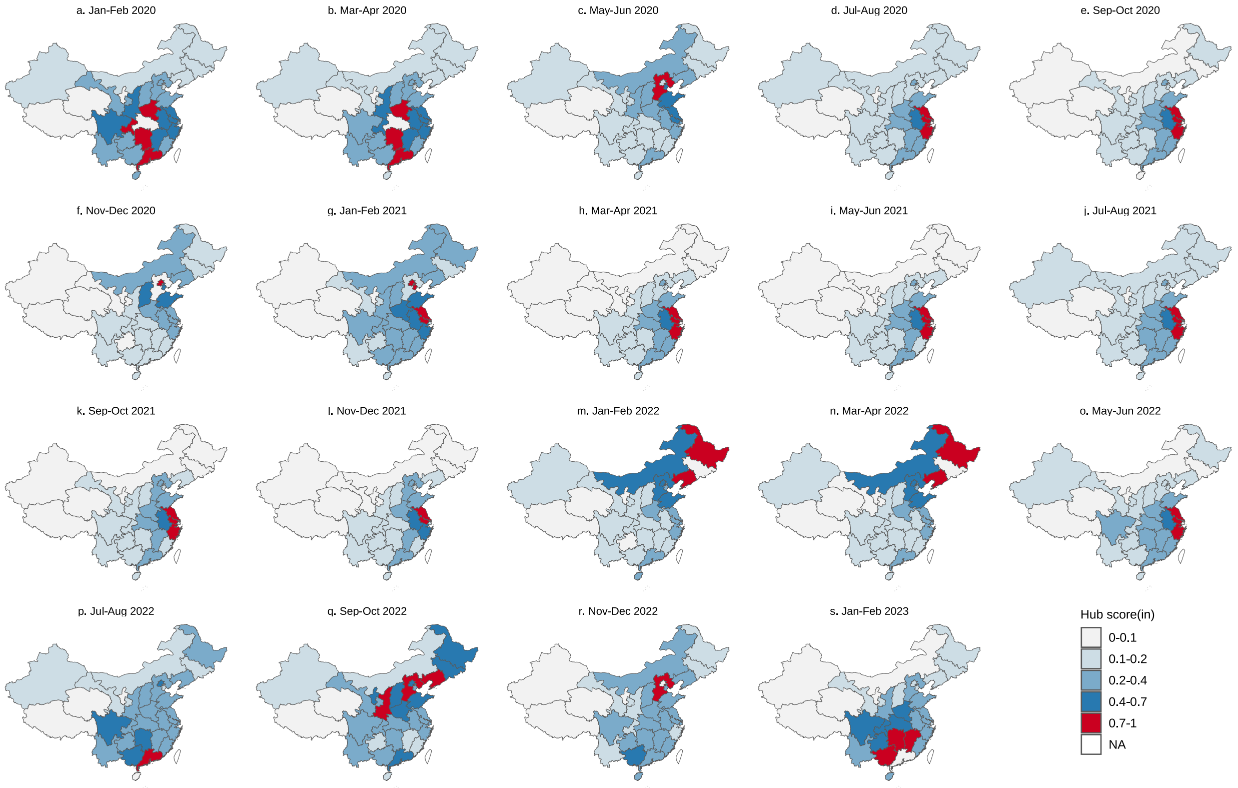


**Figure S12**. The spatiotemporal distribution of hub-score-based vital sink locations in mainland China from January 2020 to February 2023 on a bimonthly scale. The color from light to dark indicates the importance of location from weak to strong, and the importance index in specific regions is shown in the legend.

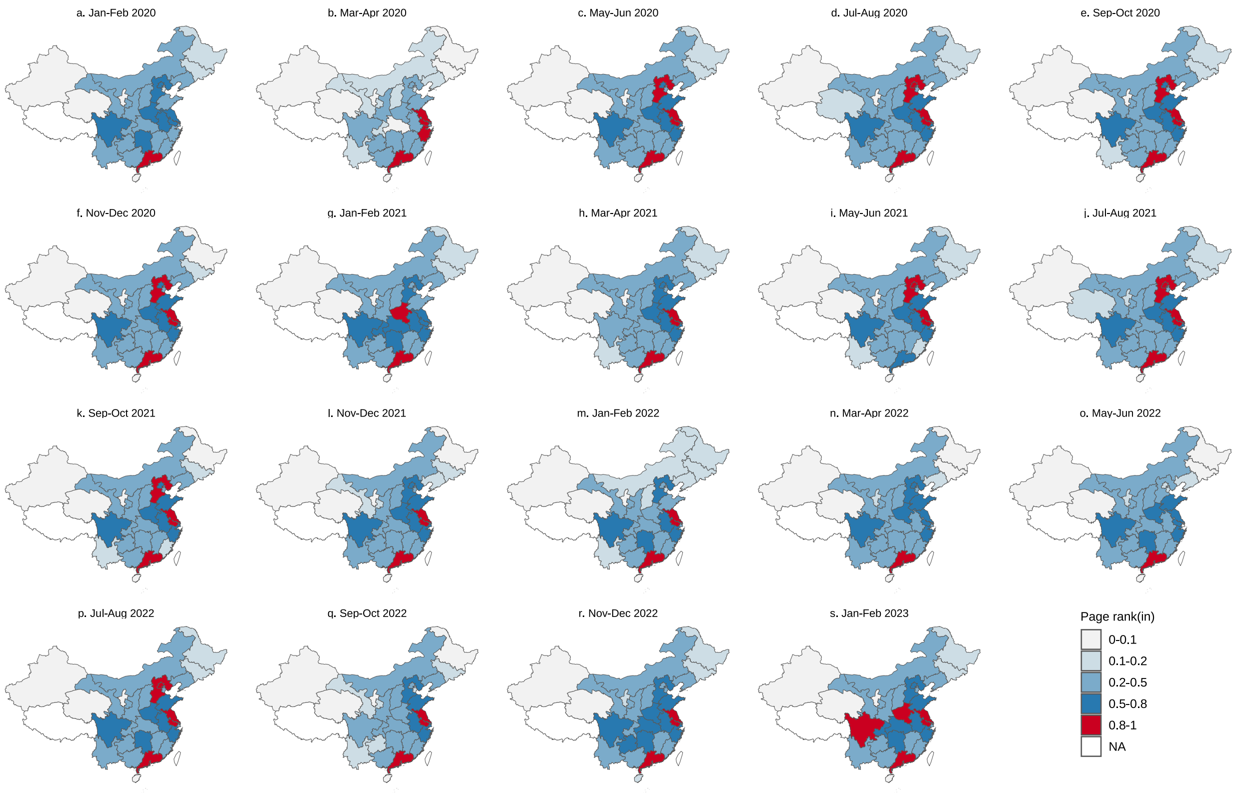


**Figure S13**. The spatiotemporal distribution of page-rank-based vital sink locations in mainland China from January 2020 to February 2023 on a bimonthly scale. The color from light to dark indicates the importance of location from weak to strong, and the importance index in specific regions is shown in the legend.

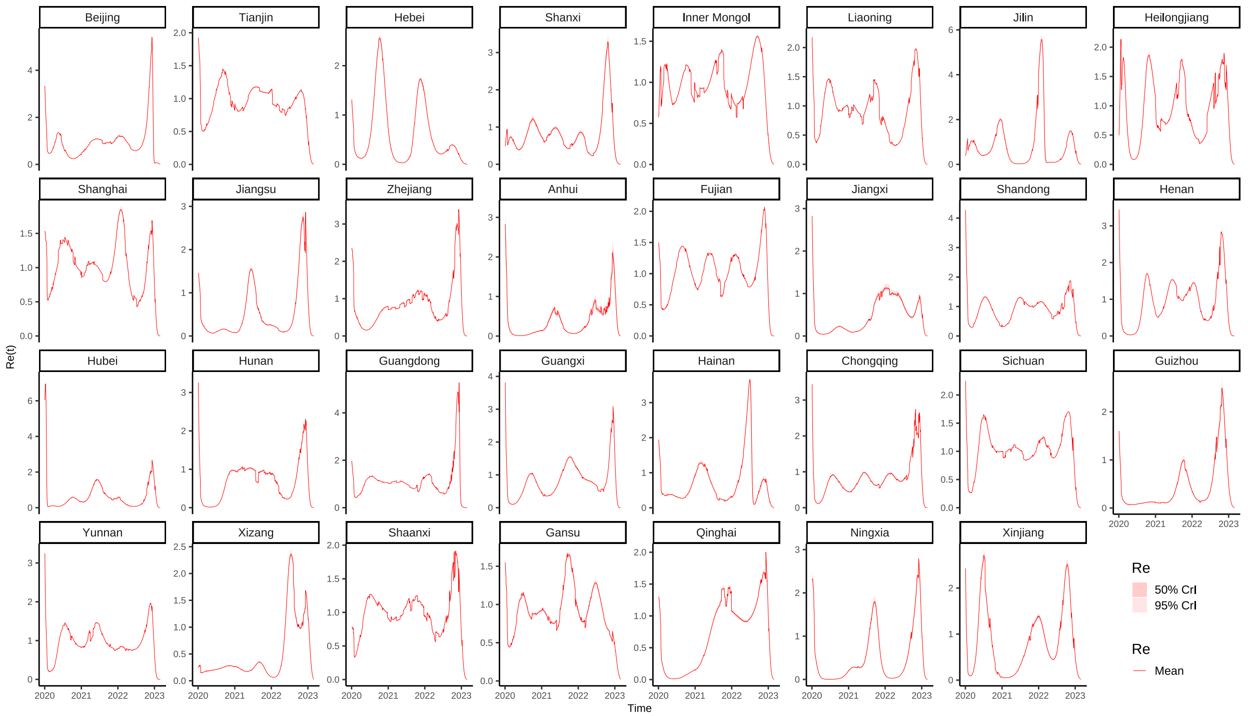


**Figure S14**. Estimated regional effective reproductive numbers across mainland China from January 2020 to February 2023, presented at a bimonthly resolution. Each panel corresponds to a different province.

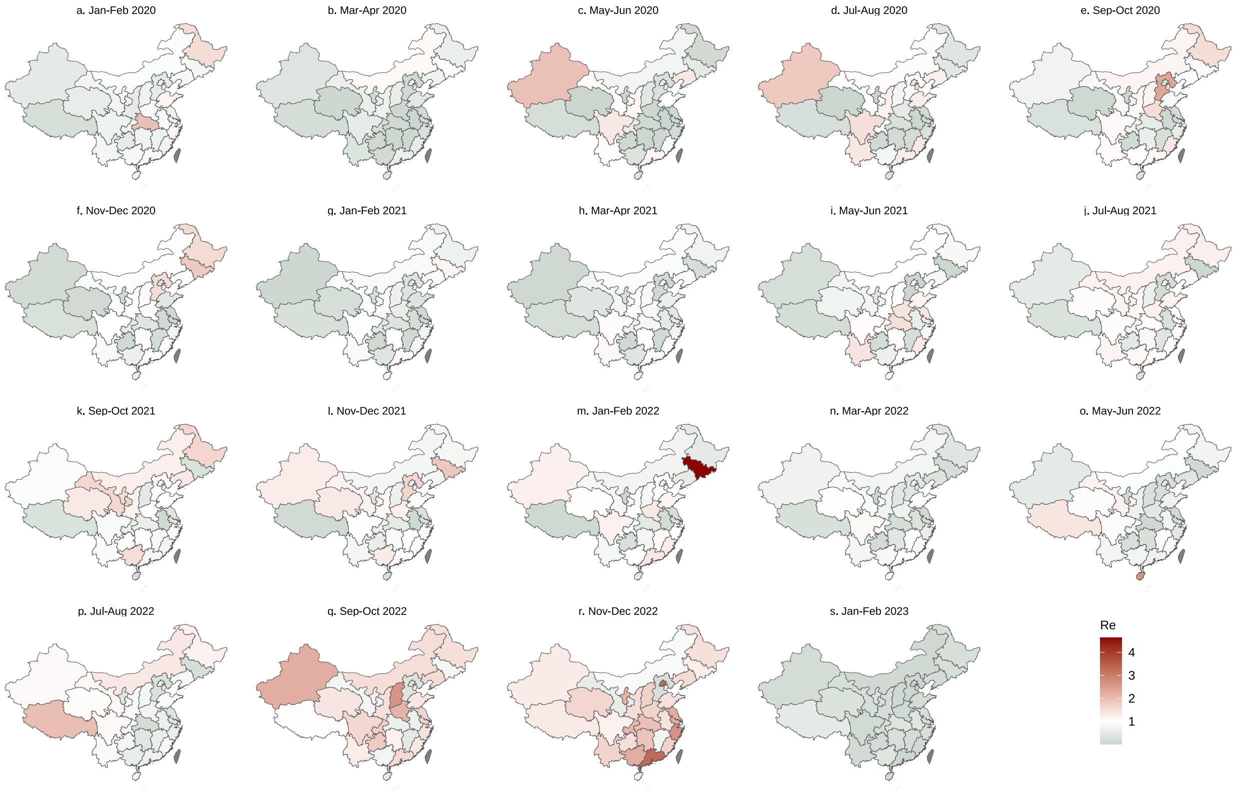


**Figure S15**. The spatiotemporal distribution of the effective reproductive number in mainland China from January 2020 to February 2023 on a bimonthly scale. The color from light to dark indicates the rates from less to high.

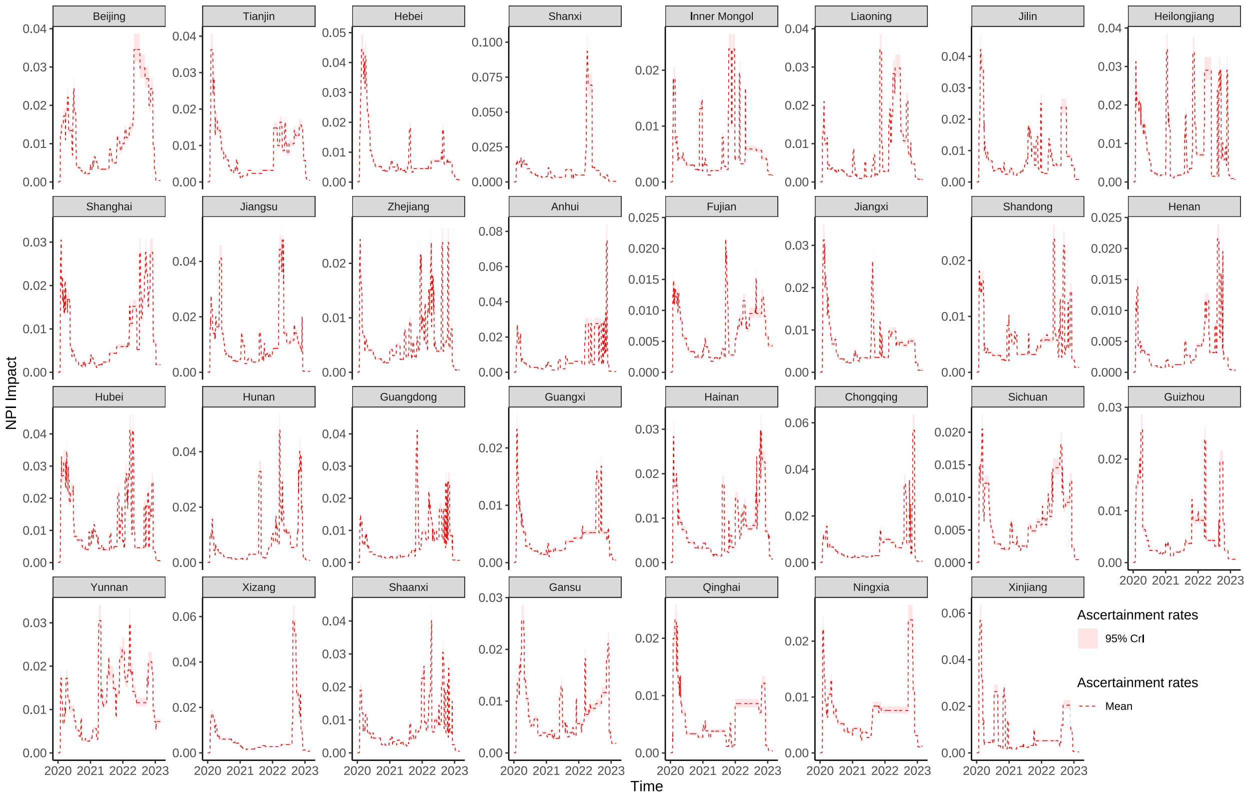


**Figure S16**. Bimonthly estimates of regional ascertainment rates across mainland China from January 2020 to February 2023. Each panel depicts data corresponding to an individual province.

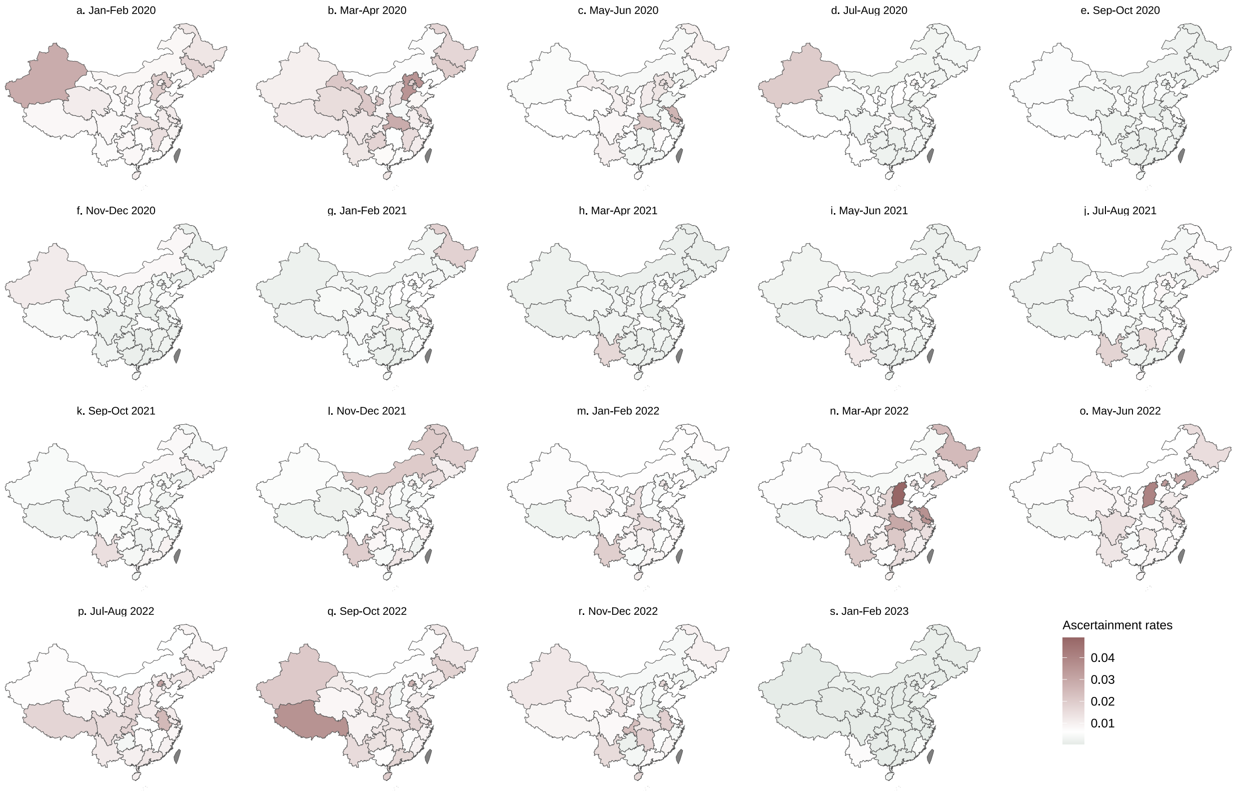


**Figure S17**. Spatiotemporal distribution of ascertainment rates in mainland China from January 2020 to February 2023, resolved at a bimonthly scale. Shading from light to dark represents increasing ascertainment rates, with specific regional values detailed in the legend.
